# Effects of the ActiveBrains trial on cardiometabolic and mental health in children with overweight or obesity: A Randomized Clinical Trial

**DOI:** 10.1101/2022.07.26.22278050

**Authors:** Jairo H Migueles, Cristina Cadenas-Sanchez, David Revalds Lubans, Pontus Henriksson, Lucia V Torres-Lopez, María Rodriguez-Ayllon, Abel Plaza-Florido, José J. Gil-Cosano, Hanna Henriksson, María Victoria Escolano-Margarit, José Gómez-Vida, José Maldonado, Marie Löf, Jonatan R Ruiz, Idoia Labayen, Francisco B Ortega

## Abstract

**Importance:** Childhood obesity is a risk factor for type 2 diabetes (T2D), cardiovascular disease, and mental disorders later in life. To investigate the parallel effects on cardiometabolic and mental health in children with overweight or obesity will provide new insights on the benefits of exercise on overall health.

**Objective:** This study aimed to investigate the effects of a 20-week exercise program on cardiometabolic and mental health in children with overweight or obesity.

**Design:** Parallel-group randomized clinical trial (RCT) conducted in Granada (Spain) from November 2014 to June 2016.

**Setting:** Clinical setting.

**Participants:** Eligibility criteria included children with overweight or obesity aged 8 to 11.9 years from Granada (Spain) and surrounding areas.

**Intervention:** The exercise program included 3-5 sessions/week (90 min/session) of aerobic plus resistance training for 20-weeks. The wait-list control group continued with their usual routines.

**Main Outcomes and Measures:** Cardiometabolic outcomes included body composition (fat mass, fat-free mass, and visceral adipose tissue), physical fitness (cardiorespiratory, speed-agility, and muscular), and traditional risk factors (waist circumference, blood lipids biomarkers, glucose, insulin, and blood pressure). Cardiometabolic risk score (z-score) was calculated based on age and sex reference values for triglycerides, inverted high-density lipoprotein cholesterol, glucose, the average of systolic and diastolic blood pressure, and waist circumference. An additional cardiometabolic risk score also included cardiorespiratory fitness. Mental health outcomes included an array of psychological well-being and ill-being indicators.

**Results:** The ActiveBrains exercise program reduced the cardiometabolic risk score by ∼0.4 (95% confidence interval [CI_95%_]: −0.75, −0.03) standard deviations (SD). The exercise program had a positive effect on low-density lipoprotein cholesterol (−7.40 [CI_95%_: −14.82, 0.016] mg/dL), body mass index (−0.60 [CI_95%_: −1.07, −0.13] kg/m^2^), fat mass index (−0.70 [CI_95%_: −1.03 to −0.36] kg/m^2^), visceral adipose tissue (−34.05 [CI_95%_: −61.38, −6.73] g), and cardiorespiratory fitness (+3.07 [CI_95%_: 0.68, 5.45] laps) in the exercise group compared to controls. No effects were observed on mental health outcomes.

**Conclusions and Relevance:** The ActiveBrains exercise program improved cardiometabolic health in children with overweight or obesity, yet it had no effect on mental health. These findings support public health initiatives promoting exercise programs in children with excess body weight to prevent future cardiovascular comorbidities.

**Trial Registration:** ClinicalTrials.gov Identifier: NCT02295072.

**Key Points:** *Question:* What are the parallel effects of exercise on cardiometabolic and mental health in children with excess of adiposity?

*Findings:* In this parallel-group randomized clinical trial of 109 children with overweight or obesity, a 20-week exercise program including aerobic plus resistance training improved body composition, cardiorespiratory fitness, and cardiometabolic risk factors. No effects on mental health were observed.

*Meaning:* Exercise programs should be promoted in children with excess adiposity to improve their cardiometabolic health.

## Introduction

Obesity is a major risk factor for type 2 diabetes (T2D) and cardiovascular disease (CVD),^1^ and there has been alarming increase in the prevalence of T2D among youth with obesity.^2,3^ Best practice prevention of T2D and CVD should initiate with fighting against obesity in childhood. Other comorbidities associated with pediatric obesity include poor cardiometabolic^4–6^ and mental health.^7^ Exercise is considered an essential component of obesity treatment programs in children due to its physical, psychological, and cognitive benefits.^8^

Previous trials in children with obesity have demonstrated exercise-induced improvements in visceral fat,^9,10^ high- and low-density cholesterol (HDL, LDL),^9,11^ insulin resistance,^10^ blood pressure,^12^ body composition,^10,11^ cardiorespiratory fitness (CRF),^10–12^ and psychological well-being.^13^ Effects have varied across studies, and none of these trials have included a cardiometabolic risk score as an outcome.^14,15^ A recent scoping review on the topic stated that measuring cardiometabolic risk as a composite or clustered score that includes measures of adiposity, lipids, metabolism, and blood pressure at childhood is a better predictor of CVD in young adulthood than other categorical measures (e.g., the presence of metabolic syndrome).^16^ Likewise, cardiometabolic risk scores have proven to be a better marker of cardiovascular health in children than single risk factors.^17^ In adolescents with obesity, the HEARTY study demonstrated exercise benefits for cardiometabolic health,^18^ physical fitness,^19^ and mental health;^20^ yet these parallel effects have not been ever demonstrated in younger children as noted in a recent systematic review and meta-analysis.^21^

A recent consensus statement called the attention on the relevance of exploring and understanding the exercise response variability,^22^ yet information is limited on the individual variability of exercise effects in children with obesity.^23^ Therefore, the primary aim of this study was to investigate the effects of a 20-week exercise program on cardiometabolic and mental health in children with overweight or obesity. Our secondary aim was to examine the within-individual variability in the effects observed.

## Methods

### Study design

A randomized clinical trial (RCT, Clinical Trial registration no. NCT02295072) to investigate the effects of exercise on brain and cognitive function in children with overweight and obesity.^24,25^ The trial protocol can be found in Supplement 1. Consolidated Standards of Reporting Trials (CONSORT) guidelines were followed for the reporting of trial results (Supplement 2).^26^ This study presents the effects on secondary outcomes from the ActiveBrains, the primary outcomes study and main effects can be found elsewhere.^24^

### Ethics

The ActiveBrains RCT was approved by the Human Research Ethics Committee of the University of Granada. Written informed consent was obtained by the parents or the legal guardians of all participants, who provided informed assent.

### Participants

Pre-pubertal children (8-11 years) with overweight or obesity, and not presenting any neuropsychological (including attention-deficit hyperactivity disorder) or physical problems were eligible to participate in the ActiveBrains RCT (see Supplement 1).

### Randomization and masking

Participants were randomly assigned to either the exercise program or the wait-list control group with simple random allocation in a ratio of 1:1 by a blinded individual not involved in the exercise sessions or the outcome evaluations (FBO). Randomization was performed immediately after the baseline evaluation, and the physical trainers running the exercise program were not involved in the outcome evaluations or randomization.

### Procedures and Interventions

The ActiveBrains exercise program had a duration of 20 weeks and was based on the global physical activity recommendations for children, including aerobic and muscle-bone strengthening activities (hereafter referred to as resistance exercise).^27^ The exercise group were instructed to attend at least three (out of five offered) supervised sessions/week. Each session lasted 90 min (60 min of aerobic plus 30 min of resistance exercises). Heart rate monitors (POLAR RS300X, Polar Electro Oy Inc., Kempele, Finland) were used to track participants’ exercise intensity during sessions. Children spent an average of 38 min per session above 80% of their HR_max_. Participants in the control group continued with their usual routines. Both control and exercise groups were provided with information about healthy nutrition and physical activity recommendations at the beginning of the study. Detailed information on the exercise program can be found in **Supplement 1**.

### Outcome measures

Measurements were conducted at baseline and immediately after the end of the intervention. Sociodemographic data were reported by children and their parents. At baseline, somatic maturation was assessed with the peak height velocity from height and sitting height measurements using the Moore’s equations.^28^

#### Cardiometabolic health

Cardiometabolic health outcomes included traditional risk factors for cardiometabolic risk score (i.e., hyperglycemia, hypertension, and dyslipidemia),^25^ as well as body composition, and physical fitness, which are closely related to cardiometabolic health.^29,30^ Blood lipids biomarkers included fasting LDL- and HDL-cholesterol, and triglycerides. The triglycerides-to-HDL ratio was calculated. Fasting insulin and glucose were obtained from blood samples and the homeostatic model assessment (HOMA) index was calculated as insulin (μU/mL) multiplied by glucose (mg/dL) and divided by 405. All blood samples were collected at the hospital between 8:00 am and 10:30 am after a minimum of 8h overnight fasting and were analyzed by an accredited laboratory. Systolic and diastolic blood pressure were assessed twice in a sitting position from the left arm with an automatic sphygmomanometer (Omron M6, Hoofddorp, The Netherlands), and the lowest values out of the two measures was considered. The systolic and diastolic average, and the mean arterial pressure were calculated. Then, the risk of dyslipidemia (based on an alteration of triglycerides and/or HDL), pre-diabetes (glucose), and pre-hypertension (systolic and diastolic blood pressure) were classified based on the age- and sex-specific cut-offs that are linked to the Adult Treatment Panel III and International Diabetes Federation criteria.^31^

Body weight and height were measured twice using an electronic scale and a stadiometer (SECA, Hamburg, Germany). Body mass index was calculated as kg/m^2^ and used to derive the age- and sex-specific z-scores according to the World Health Organization references. Whole-body fat mass and lean mass, and visceral adipose tissue were measured via dual-energy X-ray absorptiometry (DXA, Discovery Horizon® DXA system, Hologic, Canada ULC). Fat mass index and lean mass index were calculated (kg/m^2^). Abdominal obesity was represented by the average waist circumference from two measurements.^32^ Physical fitness components CRF, speed-agility, and muscular fitness were assessed using the 20 m shuttle-run (laps and estimated VO_2_max^33,34^), the 4×10 m shuttle run (time to complete the circuit), and the handgrip (kg) and standing long jump (cm) tests, respectively. These tests are valid, reliable, and feasible.^35–37^ Detailed information on the physical fitness testing can be found elsewhere.^25^ Then, unfit children were classified based on age- and sex-specific international reference values for cardiorespiratory fitness in children.^38^

Finally, a previously validated cardiometabolic risk score was calculated.^14^ The score averaged the age-and sex-specific z-scores (based on European reference values)^15^ for triglycerides, inverted HDL, glucose, and the average of systolic, and diastolic blood pressure, and waist circumference.^14^ Since the American Heart Association has recently proposed CRF as powerful marker cardiometabolic health,^39^ we additionally included CRF performance in a cardiometabolic risk score 2. Based on European reference values, children with at risk of metabolic syndrome were identified as those with a z-score ≥0.39 (i.e., deviating 0.39 standard deviations [SD] from the European pediatric population) as previously proposed.^15^

#### Mental health

Children completed the mental health questionnaires on three separate days. Both psychological ill-being and well-being components of mental health were assessed using valid self-reported questionnaires. Psychological ill-being measures included stress (Children’s Daily Stress Inventory, scored from 0 to 30), anxiety (State-Trait Anxiety Inventory for Children, scored from 20 to 60), depression (Children’s Depression Inventory, scored from 0 to 54) and negative affect (Positive and Negative Affect Schedule for Children, scored from 10 to 30). Psychological well-being measures included positive affect (Positive and Negative Affect Schedule for Children, scored from 10 to 30), happiness (Subjective Happiness Scale, scored from 4 to 28), optimism (Life Orientation Test-Revised, scored from 6 to 30), self-efficacy (General Self-Efficacy, scored from 10 to 40), self-concept (Five-Factor Self-concept questionnaire, scored from 30 to 300) and self-esteem (Rosenberg Self-Esteem Scale, scored from 10 to 40). A detailed description of the mental health indicators assessment and the tests psychometric information can be found elsewhere.^25^ Composite standardized scores were calculated for psychological ill-being (i.e., stress, anxiety, depression and negative affect), psychological well-being (i.e., positive affect, happiness, optimism, self-efficacy, self-concept, and self-esteem) and total mental health (i.e., psychological ill-being multiplied by −1 and psychological well-being). Additionally, we also calculated the risk of anxiety (cut-off score ≥ 40 for state anxiety)^40^ and depression (cut-off score ≥ 19)^41^.

#### Physical activity assessment

Accelerometer-determined daily time spent in physical activity, sedentary behavior (SB), and sleep during the intervention were used to assess the change in daily activity induced by the exercise intervention. Accelerometers (GT3X+, ActiGraph, Pensacola, Florida, USA) were placed on the right hip and the non-dominant wrist to monitor physical activity for seven days at baseline and during the intervention delivered period for exercise and control groups. The accelerometers raw data were processed as described elsewhere,^42^ following the practical recommendations previously done by our group.^43^ In brief, a minimum of four valid days (i.e., ≥16 hours/day), including at least one weekend day, was required to be included in the analyses. We used the GGIR software^44^ to identify the night sleep periods using an automated algorithm guided by the self-reported sleep times.^45,46^ Waking time was classified into MVPA, light physical activity (LPA), and SB using children-specific cut-points.^47–49^

### Sample size calculation

A posteriori power analyses showed that a sample size of 98 children is enough to detect small-to-medium effect sizes (i.e., 0.3 SDs) assuming an α error of 0.05 and 80% statistical power.

### Statistical analysis

Characteristics of the study participants are presented as mean and SD, or frequency and percentage. Prior to analyses, raw scores from each outcome were winsorized (when needed) to limit the influence of extreme values.^50^ Then, baseline z-scores of the outcomes were calculated by subtracting their mean and dividing by their SD. Post-exercise z-scores were calculated relative to the baseline mean and SD as a standardized measure of the effect size.^50^ Analysis of covariance (ANCOVA) models were built including post-exercise outcome values as dependent variables, group (i.e., exercise vs. control) as fixed factor, and baseline levels of the outcome studied as covariate.^50^ Analyses were primarily conducted under the per-protocol principle, i.e., attending to 70% of the sessions, and sensitivity analysis were performed under the intention-to-treat principle including all participants and imputing the missing data using predictive mean matching multiple imputations.^51^ We did not observe strong evidence supporting that missing data was not missing at random. The within-individual change distribution was studied and the changes exceeding 0.2 Cohen’s D were considered meaningful (accepted threshold for relevant standardized effect size).^23^ Chi-square tests were used to compare the rate of meaningful changes observed in the exercise and the control group. Additionally, we explored the change in the daily distribution of the movement behaviors induced by the exercise program. This comparison was performed following the compositional data analysis standards,^52^ in line with the conclusions reported a previous expert consensus for the analysis of device-measured movement behaviors.^53^ Values in the change composition are represented as proportional changes (%) from the baseline overall composition, and the Hotelling’s T-squared test for multivariate pair-wise comparisons was used. All the statistical procedures were performed using the R software (v. 4.0.0.). A significant difference level of *P*<0.05 was set.

## Results

Out of the 109 participants enrolled, 98 were included in the per-protocol analysis (**Figure 1**). Participants’ characteristics are presented in **eTable 1** in **Supplement 3**. At baseline, 43% of the children were at risk of dyslipidemia, 3% presented pre-diabetes, 9% pre-hypertension, 77% were unfit, 24% at risk of metabolic syndrome, 19% at risk of anxiety, and 3% at risk of depression. No significant differences were found regarding baseline characteristics (e.g., age, body mass index, fat mass index, parental educational level) between compliant participants (n=98), and the rest of the participants measured at baseline (n=11, all *P*>0.05).

**Figure 1.**
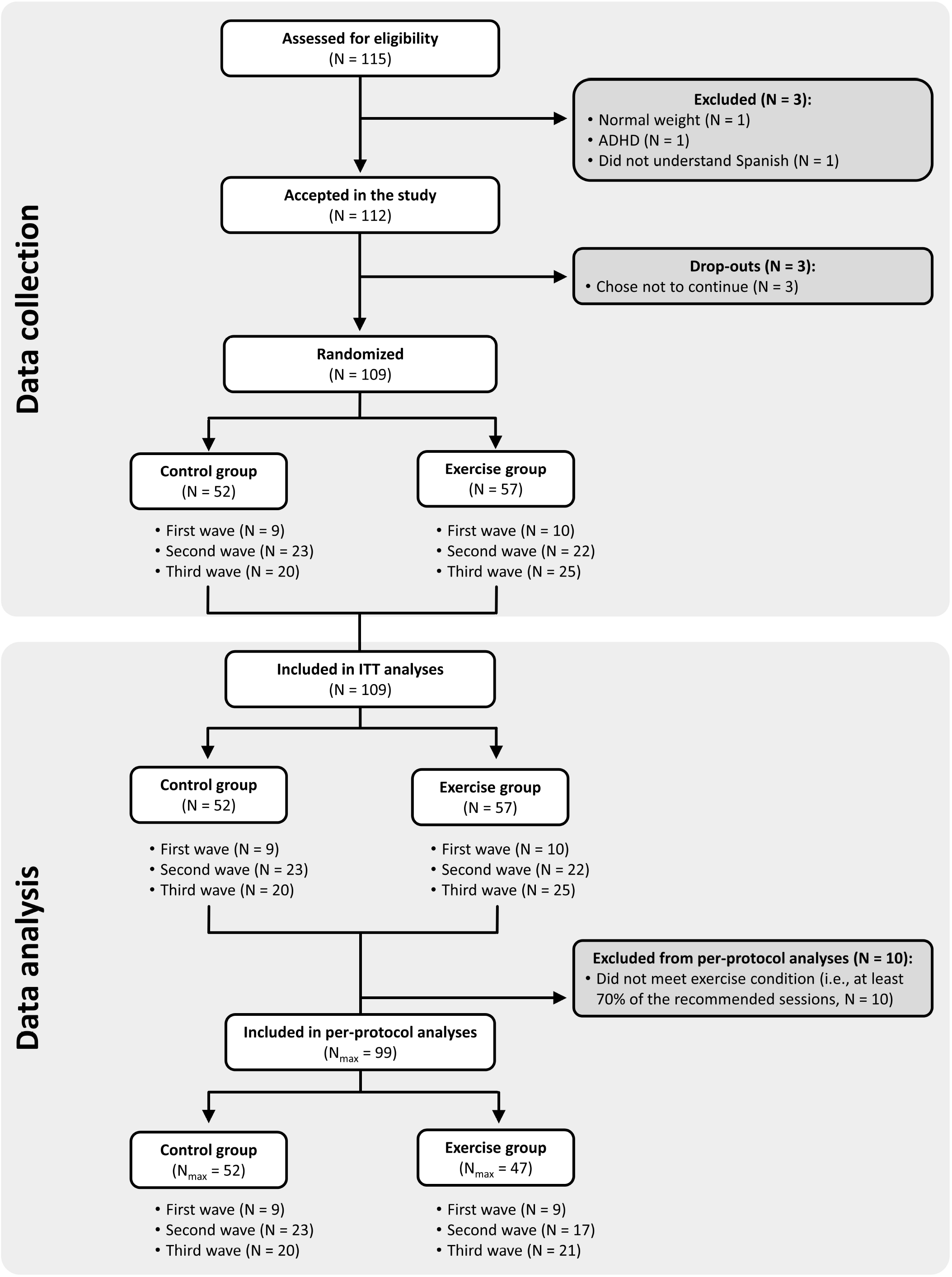
Flowchart of the study. ADHD: Attention-deficit hyperactivity disorder; ITT: Intention-to-treat. For final ITT analyses, those participants that left the study during the exercise program or did not complete the post-exercise program assessments were imputed (see Statistical section). N_max_: Maximum N for analyses, it changes depending on the variable, see eTable 2 to 5 in Supplement 3 for the main study outcomes.

### Cardiometabolic health

Figure 2. shows the within- and between-groups pre-post differences in cardiometabolic health outcomes (tabulated data in **eTable 2** in **Supplement 3**). The ActiveBrains exercise program produced a meaningful reduction in cardiometabolic risk (score 1: −0.36 [-0.72, 0.00] SDs; score 2: −0.38 [-0.74, −0.02] SDs). We found a borderline non-significant reduction in LDL of 7.0 (95% confidence interval [CI_95%_]: −14.3, 0.4) mg/dL, and significant reductions in body mass index (−0.59 [CI_95%_: −1.06, −0.12] kg/m^2^), fat mass index (−0.67 [CI_95%_: −1.01 to - 0.33] kg/m^2^), and visceral adipose tissue (−31.4 [CI_95%_: −58.99, −3.90] g) in the exercise group compared to the control group. The exercise group improved CRF performance (+2.75 [CI_95%_: 0.22, 5.28] laps) and estimated VO_2_max (+0.94 [0.05, 1.84] ml/kg/min) compared to the control group. Overall, the intention-to-treat analyses showed similar but attenuated effects (**eTable 3** in **Supplement 3**).

**Figure 2.**
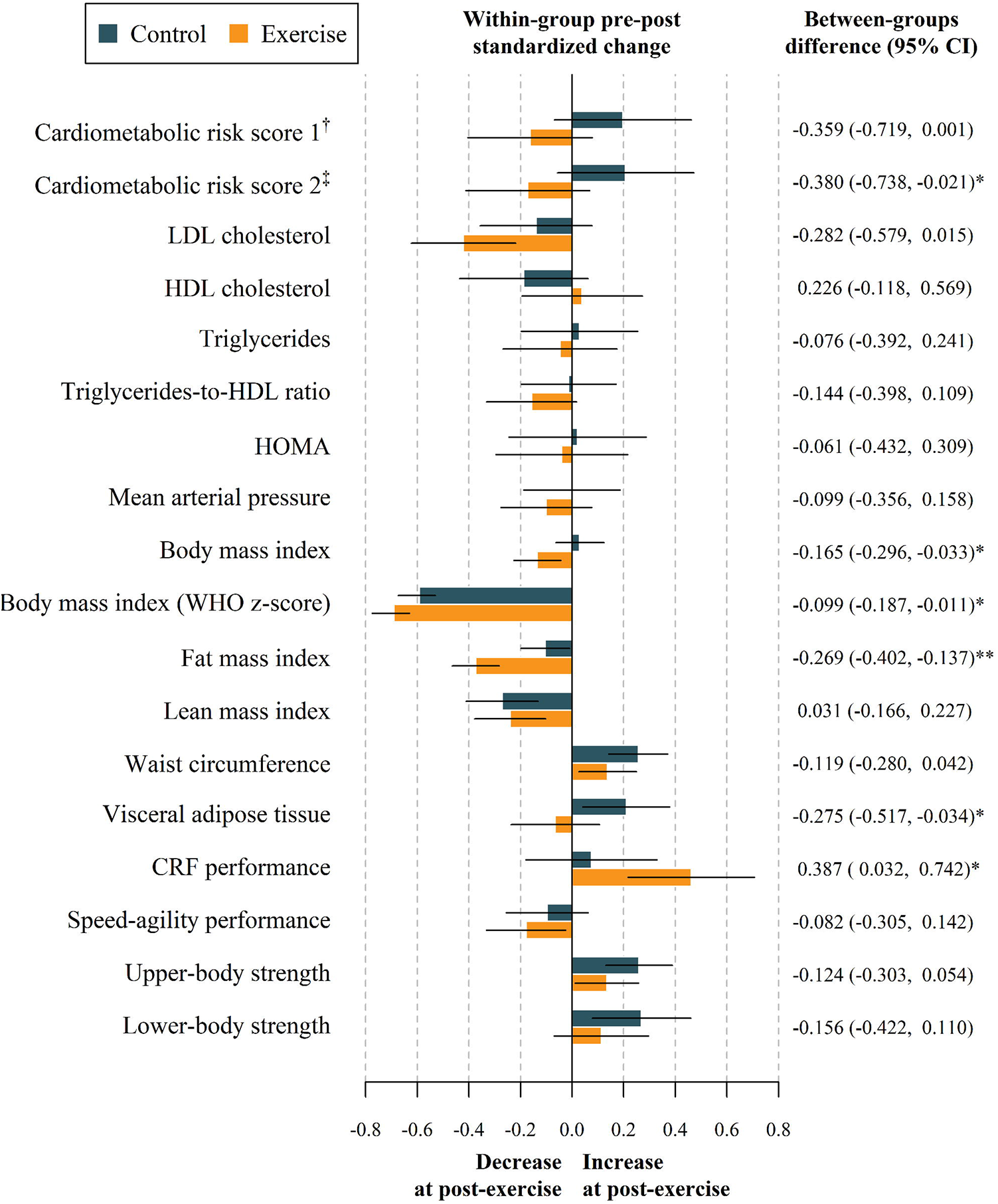
Effects of the ActiveBrains exercise program z-score pre-post change between groups in cardiometabolic risk outcomes (per-protocol analyses). Data analyses were primarily conducted under the per-protocol principle, i.e., attending to 70% of the sessions. Baseline z-score of the outcomes were calculated by subtracting the mean value and dividing by the SD of each outcome. Post-exercise z-scores were calculated relative to the mean and SD of the baseline values, being a z-score of the change in each outcome, i.e., (post-exercise_*i*_ – baseline mean) / baseline SD. ^†^Cardiometabolic risk score 1 was calculated as the age- and sex-normalized scores for HDL cholesterol, waist circumference, triglycerides, glucose, and the average of systolic and diastolic blood pressure based on European population reference values. ^‡^Cardiometabolic risk score 2 additionally included the CRF. LDL: low density lipoprotein cholesterol; HDL: high density lipoprotein cholesterol; HOMA: homeostatic model assessment; CRF: cardiorespiratory fitness as measure by laps in the 20m shuttle run test \**P* < 0.05 \*\**P* < 0.01

More participants in the exercise group showed meaningful changes (i.e., ≥0.2 SDs) than in the control group in fat mass index (79% vs. 36%, *P*<0.001) and CRF performance (65% vs. 38%, *P*=0.027) **(Figure 3)**. A marginal non-significant difference was found in favor to exercise in body mass index (34% vs. 16%, *P*=0.071). Likewise, we observed that more children at risk of metabolic syndrome at baseline were not at risk after the exercise program in the exercise group compared to the control group (**Figure 4A**), and a similar trend was observed in children passing from unfit to fit status based on cardiorespiratory fitness (20% vs 5%, **Figure 4B**).

**Figure 3.**
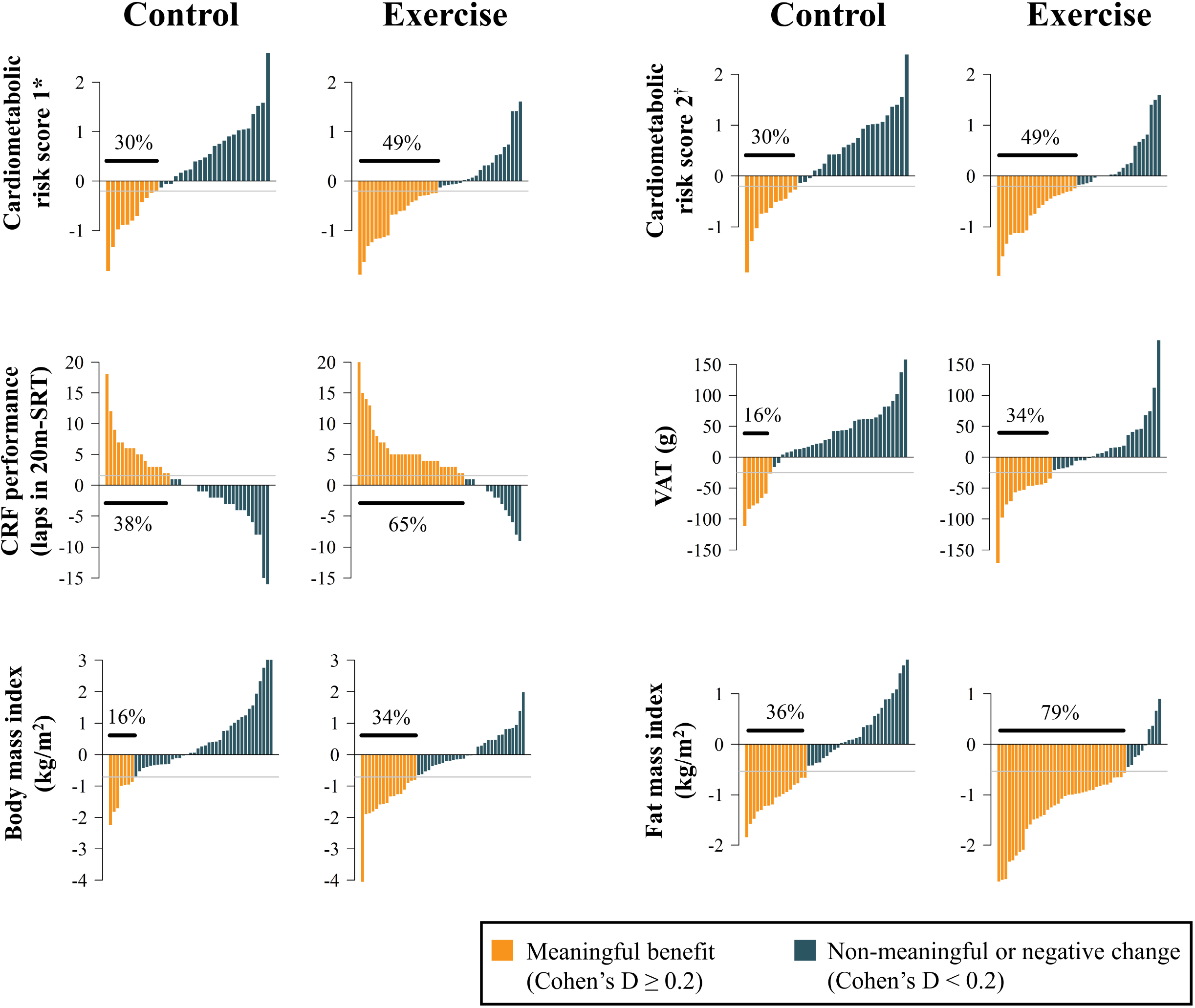
Individual change distribution in the outcomes significantly affected by the exercise program. Data analyses were primarily conducted under the per-protocol principle, i.e., attending to 70% of the sessions. *Cardiometabolic risk score 1 was calculated as the age- and sex-normalized scores for HDL cholesterol, waist circumference, triglycerides, glucose, and the average of systolic and diastolic blood pressure based on European population reference values.^15^ ^†^Cardiometabolic risk score 2 additionally included the CRF. CRF: cardiorespiratory fitness; 20m-SRT: 20m shuttle run test.

**Figure 4.**
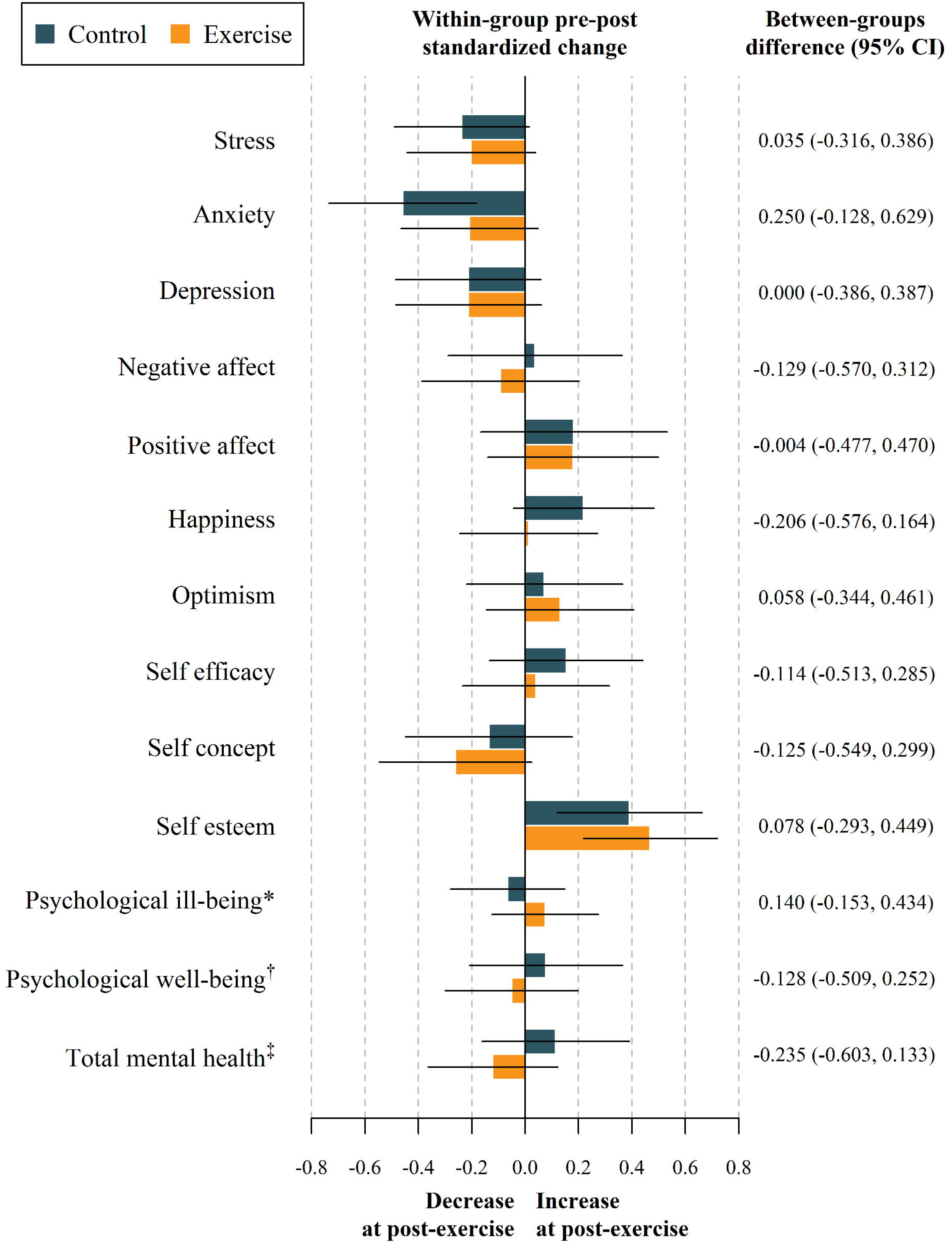
Participant rate fluctuations for (A) risk of metabolic syndrome or (B) fitness status from baseline to post-exercise. Risk of metabolic syndrome was categorized based on the average of age-and sex-specific z-scores for triglycerides, inverted high-density lipoprotein, glucose, and the average of systolic and diastolic blood pressure, and waist circumference. Based on European reference values, ^15^ those children with a z-score ≥0.39, were considered at risk of metabolic syndrome. Unfit children were classified based on age- and sex-specific international reference values for cardiorespiratory fitness in children.^38^

### Mental health

Figure 5 shows that the exercise program did not affect any mental health outcome (tabulated data in **eTable 4** in **Supplement 3**). Similarly, intention-to-treat analyses showed no effects on mental health (**eTable 5** in **Supplement 3**).

**Figure 5.**
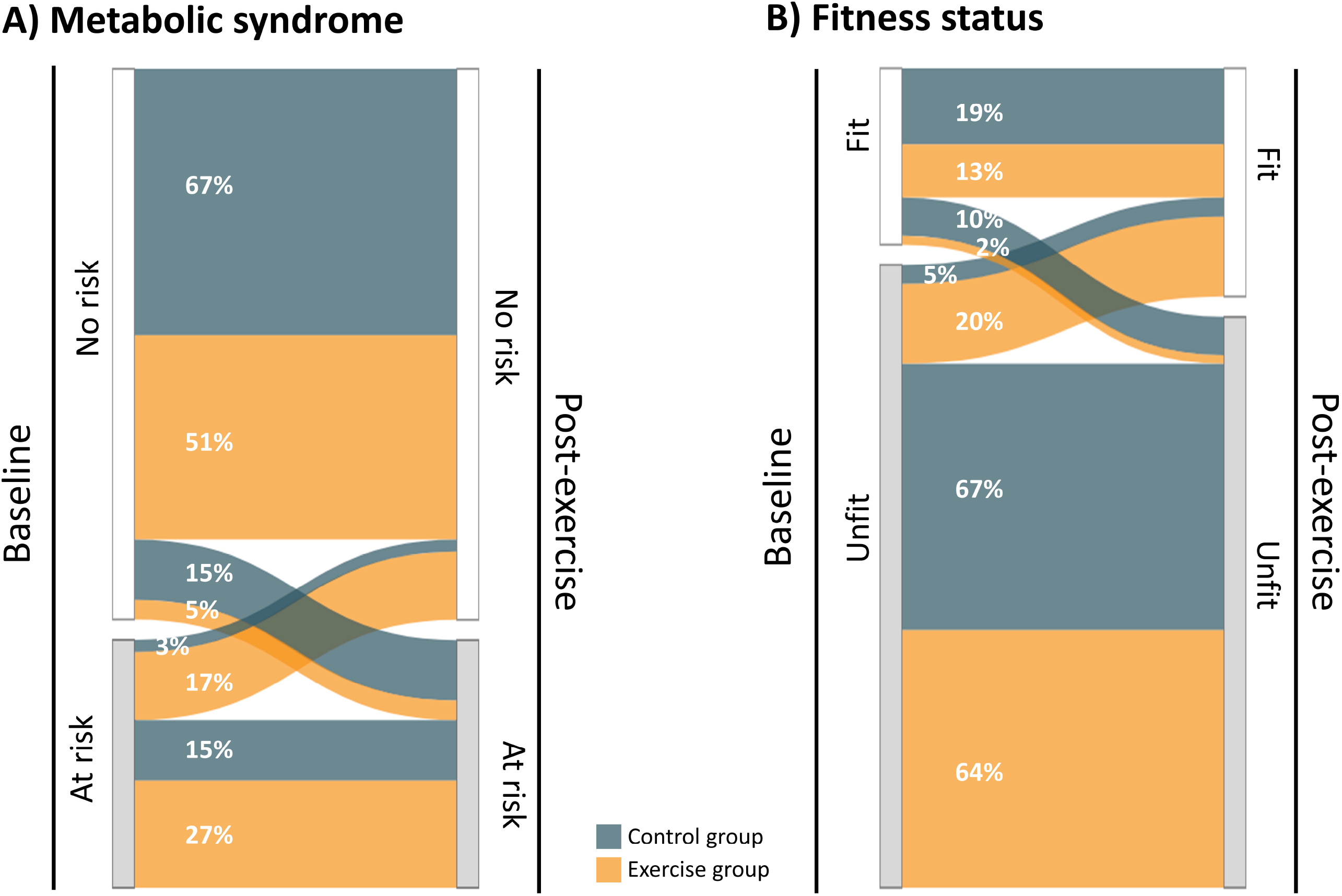
Effects of the ActiveBrains exercise program *z*-score pre-post change between groups in mental health (per-protocol analyses). Data analyses were primarily conducted under the per-protocol principle, i.e., attending to 70% of the sessions. Baseline z-score of the outcomes were calculated by subtracting the mean value and dividing by the SD of each outcome. Post-exercise z-scores were calculated relative to the mean and SD of the baseline values, being a z-score of the change in each outcome, i.e., (post-exercise_*i*_ – baseline mean) / baseline SD. *Psychological ill-being was calculated as the normalized mean of the z-score for stress, anxiety, depression, and negative affect. ^†^Psychological well-being was calculated as the normalized mean of the z-score for positive affect, happiness, optimism, self-efficacy, self-concept, and self-esteem. ^‡^Total mental health was calculated as the normalized mean of the z-score for all mental health indicators.

### Exploratory analysis: change in daily activity composition

**eFigure 1** (**Supplement 3**) shows the exercise-induced changes in physical behaviors derived from the hip- and the wrist-worn accelerometers (Panel A and Panel B, respectively). Both the hip- and the wrist-based estimations showed consistent trends that the exercise group increased MVPA compared to the control group (hip: +15% vs. +7% from baseline; wrist: +21% vs. +7% from baseline). More specifically, the wrist worn estimates resulted in a significant group-by-time effect, (*P*=0.002, Panel B), while the hip worn estimate was borderline significant (*P*=0.079, Panel A). Likewise, the control group did not substantially alter their time in LPA, SB, and sleep from baseline, while the exercise group substantially reduced their SB (hip: −6%; wrist: −14%) and sleep time (hip: −8%; wrist: −9%).

## Discussion

The ActiveBrains RCT demonstrated that a 20-week exercise program reduced the cardiometabolic risk score (composed of triglycerides, HDL, glucose, blood pressure and waist circumference; and additionally included CRF) in children with overweight or obesity. These findings were confirmed using two valid cardiometabolic risk scores.^14,39^ Specifically, the ActiveBrains exercise program substantially improved children’s blood lipids, body composition, and CRF, i.e., the individual risk factors, compared to those randomized to the control group. The proportion of children experiencing meaningful changes in cardiometabolic risk score, body composition and CRF was higher in the exercise group, compared to the control group. No significant effects were observed for the different mental health outcomes.

### Cardiometabolic health

Our study has demonstrated a sizeable reduction in cardiometabolic risk (∼0.4 SDs), and the within-individual change showed more participants at risk of metabolic syndrome at baseline were no longer at risk at post-exercise compared to controls. It seems that the risk reduction was mainly driven by improvements in blood lipids, total and visceral adiposity, and CRF, which were the cardiometabolic outcomes affected by the ActiveBrains exercise program. In agreement with the American Heart Association,^39^ our study support the use of CRF as a cardiometabolic risk factor. In addition, the exercise group reduced their fasting LDL by 7 mg/dL (i.e., 0.3 SDs) and their visceral adipose tissue by 34 g compared to the control group. Other blood lipid and adiposity markers showed a better trend in the exercise compared to the control group, yet it did not reach statistical significance (e.g., waist circumference and HDL).

Our results are consistent with recent meta-analyses in children with overweight or obesity showing that concurrent aerobic and resistance training can improve blood lipid levels, mainly LDL and triglycerides.^54,55^ Our findings are also consistent with previous research showing reductions in visceral fat,^9,10^ LDL,^9^ and increments in HDL following exercise in children with overweight or obesity.^11,12^ Two of the previous RCTs in children with obesity additionally found effects on insulin resistance,^10,12^ yet we did not. We believe that differences in the participants’ baseline characteristics may account for our lack of effects on glucose metabolism biomarkers. For example, the PLAY RCT analyzed 222 participants, of whom 28% were pre-diabetic,^10^ compared with only 3% in our study. The majority of our participants (77%) had obesity, almost 50% were at risk of dyslipidemia,^31^ 77% were unfit. Thus, there was more room for improvements in blood lipids, adiposity and CRF than there was for glycemic metabolism. Despite their weight status, our participants were at healthy glycemic and blood pressure levels at baseline, which could produce a ceiling effect.

Children in the exercise group improved their body composition by reducing their total and visceral fat mass. These results are in line with the previous literature in children with obesity regarding the reductions in body mass index and fat mass.^9–12^ Likewise, a recent network meta-analysis concluded that aerobic or the combined aerobic and resistance training effectively reduced adiposity outcomes with similar magnitude as we observed in our study (body mass index ∼0.7 kg/m^2^ vs. our findings: 0.6 kg/m^2^) in children and adolescents with overweight or obesity.^54^ No less important, we found that a higher rate of participants experienced clinically relevant change (i.e., at least 5% reduction) in their fat mass index, which is in line with the EFIGRO trial findings.^23^ Our lean mass index was not affected by the ActiveBrains exercise program, which agrees with a previous study using a similar indicator of lean mass.^9^ However, another RCT in children with obesity with a similar dose of resistance training described improvements in fat-free mass (+1.2 kg compared to controls).^12^

Regarding physical fitness, the ActiveBrains exercise program improved CRF both the performance in the test (laps) and the estimated VO_2_max. These results agree with previous trials in children with overweight or obesity.^9–12^ The ActiveBrains exercise program did not improve the children’s speed-agility or muscular fitness, which agrees with the EFIGRO trial findings, which used a similar exercise protocol in a sample of similar characteristics to ours.^9^ The specificity of our resistance exercises performed might explain this null finding, i.e., body-weight exercises instead of weightlifting may have produced benefits in muscular endurance instead of maximal strength or power (as measured by the handgrip and the standing long jump tests).

None of the previous studies have analyzed the effects of exercise programs on composite cardiometabolic risk scores, which hampers comparisons in this regard. We believe this is a strength of our study to quantify the effect on the composite cardiometabolic risk scores, which are valid measures of risk for T2D, CVD, and other cardiometabolic diseases;^14^ and better marker of cardiovascular health in children than using a single risk factor. ^17^ Our findings are further strengthened by the investigation of the proportion of children experiencing meaningful changes in the control and the exercise groups. Using a similar approach, the EFIGRO trial found higher rates of meaningful changes in hepatic fat.^23^

### Mental health

The ActiveBrains exercise program did not improve mental health (i.e., psychological ill-being and well-being) in children with overweight or obesity. The null findings for mental health might be due to the ceiling effect, i.e., most of the children had a healthy mental status at baseline. Indeed, consistent exercise effects have been observed on depression in adolescents.^21,56,57^ This is likely explained because children are still young and present high levels of well-being and low levels of ill-being, which makes unnecessary and complicated to improve these outcomes further. The effects of exercise on mental health in children and adolescents are inconsistent^57^ and differ according to a range of contextual factors (e.g., type of activity, delivery mode) and participant characteristics (e.g., age, clinical diagnosis).^58^ Seabra et al. concluded that a 20-week football program improved self-esteem in boys with overweight.^59^ Alternatively, Romero-Perez et al., found no significant changes after 20-week aerobic exercise training in anxiety and a small reduction in depression in children with obesity.^60^ Williams et al. observed that an eight-month aerobic exercise after school program provided benefits to quality of life, depressive symptoms and self-worth in children with overweight.^13^ Differences in our findings and the previous studies could be explained by the heterogeneity of the exercise program (type, only aerobic vs. aerobic and resistance training; and frequency, 2 vs. 3 to 5 sessions per week), characteristics of the study sample (sex, weight status), the mental health outcomes examined (individual dimensions vs. a complete set of psychological ill-being and well-being outcomes); and the study design (non-RCTs vs. RCT).

Although our intervention complied with most of the SAAFE principles (i.e., Supportive, Active, Autonomous, Fair, and Enjoyable) proposed by Lubans et al.^61^ to maximize the effects of exercise on mental health, we did not assess the session fidelity. Therefore, we cannot confirm that the sessions were adhered to these principles. Alternatively, the lack of sensitivity of our mental health measures and/or the ceiling effect experienced in our children may explain the null findings. Since the mental health of children with obesity is likely impaired,^2,7^ further studies proposing effective lifestyle interventions to improve their mental health are urgently needed. We recommend that future trials monitor the acute effects on mental health in every session to find the best type, frequency, intensity, and duration of exercises to target mental health.

### Strengths and limitations

The strengths of our study include: the randomized design; the holistic view of both physical and mental health with a complete array of outcomes in children with obesity; the quantification of the cardiometabolic risk; the description of the distribution of the meaningful change at individual level; the use of gold-standard measures of cardiometabolic health and body composition, the use of reliable and valid physical fitness tests and mental health questionnaires; and the heart rate monitoring using individualized and individually programmed heart rate monitors based on the previous maximal exercise test. However, there are some limitations that should be noted. These study findings might be limited by the relatively low sample size for some outcomes, which could make some of the statistical analyses underpowered to detect significant differences and by the fact that some of the evaluators were not blinded to the group allocation. We believe that most of the outcomes included in our study are objective and unlikely to be influenced by assessor blinding (i.e., cardiometabolic health, blood markers assessed in external laboratory, and body composition by DXA). However, it is possible that the lack of findings in mental health is explained by the ceiling effect observed in our children (i.e., healthy mental status at baseline).

## Conclusion

The ActiveBrains exercise program improved cardiometabolic health in children with overweight or obesity. The cardiometabolic risk score was be reduced by ∼0.4 SDs, which was mainly due to the improvements observed in blood lipids, total and visceral adiposity, and CRF. Our intervention did not however affect any of the mental health studied. These findings support public health initiatives promoting exercise programs in children with obesity have the potential to improve their cardiometabolic health. In needs to be further investigated the no effect on mental health outcomes, including whether the instruments are sensitive enough to detect changes, whether there is a ceiling effect in young children who might be mentally healthy overall, etc.

## Supporting information

Supplement 1, Trial protocol

Supplement 2, CONSORT checklist

Supplement 3 eTables and eFigures

## Data Availability

We did not obtain children parents consent to widely share the data nor was it included in the IRB protocol.

## Author Contributions

Drs Migueles and Cadenas-Sanchez had full access to all the data in the study and take responsibility for the integrity of the data and the accuracy of the data analysis. FBO, IL and JRR designed research; JHM, CCS, DRL, PH, LVTL, MRA, APF, JJC, HH, MVEM, JGV, JM, ML, JRR, IL, FBO performed research; JHM analyzed data; and JHM and CCS was in charge of drafting the manuscript. All authors critically revised the manuscript, contributed to the conception and interpretation of the analyses and approved the final version of the manuscript to submitted.

