## Supplement 1, Trial protocol for "Effects of the ActiveBrains trial on cardiometabolic and mental health in children with overweight or obesity: A Randomized Clinical Trial"

**CLINICAL TRIAL PROTOCOL**

**Effects of an exercise-based randomized controlled trial on cognition, brain structure and brain function in overweight preadolescent children (ActiveBrains)**

**Brief title**

Effects of an exercise program on cognition and brain in overweight/obese preadolescent children

**Organization’s Unique Protocol ID**

DEP2013-47540-R

**Study Start**

December 2014

**Study Completion**

July 2017

**Sponsor**

Universidad de Granada

**Principal Investigator**

Francisco B. Ortega Porcel. Professor at the University of Granada, Granada, Spain.

**Principal Collaborators**

Research Institute of Sports and Health (iMUDS), Granada, Spain

Research Center of Brain, Mind and Behavior (CIMCYC), Granada, Spain

*Biomedical Research Centre (CIBM)*, Granada, Spain

San Cecilio University Hospital, Granada, Spain

Virgen de las Nieves Hospital, Granada, Spain

**Human Subjects Review**

Approval Number: 848

Board Name: Ethics Committee on Human Research

Board Affiliation: University of Granada

### **1. PROTOCOL SUMMARY**

New advances in neuroelectric and neuroimaging technologies in the last years provide a golden opportunity to further explore and understand how cognition and brain function can be stimulated by environmental factors, such as exercise, and particularly to study whether physical activity influences brain development in early ages. The present study, namely the ActiveBrains project, aims to examine the effects of a 20-week physical exercise program on cognition and brain, as well as on selected physical and mental health outcomes in preadolescents overweight/obese children. A total of 109 children with overweight/obesity aged 8 to 11.9 years will be randomized into an exercise group and a control group. The intervention will last 20 weeks, with 3-5 after-school sessions/week of 90 min each and will focus on high-intensity aerobic exercise mainly yet also includes muscle-strengthening exercises. The extent to what the intervention effect remains 8-months after the exercise program finalizes will also be studied in a subsample. Edge-cutting technologies will be used to assess cognitive performance, brain structure and function, by means of magnetic resonance imaging examination (both structural and functional) and an electroencephalogram examination (event-related brain potentials). The secondary outcomes will be grouped as physical health outcomes (e.g., physical fitness, body fatness, and bone mass and lipid-metabolic factors) and mental health outcomes (e.g. chronic stress indicators and overall behavioral and personality measurements such as anxiety or depression). Cross-sectional and longitudinal studies support that exercise might benefit cognition/brain in children; however, well-designed randomized controlled trials are needed to confirm or contrast these observational data, and to explore the causal pathways responsible for such associations. This project will substantially contribute to the existing knowledge and will have an impact on societies, since early stimulation of brain development might have long lasting consequences on cognitive performance, academic achievement and in the prevention of behavioral problems and the promotion of mental health.

### **2. BACKGROUND**

#### 2.1. State of the art and rationale for the primary aim

Over the last years, schools in USA and in some European countries have received pressure from different entities to reduce the time devoted to physical education in the school curriculum, arguing that more time spent in traditional and standard academic tasks would result in improvements in academic achievement. Several systematic reviews consistently suggest that this is a wrong assumption. Studies focused on increasing physical education time within the school hours and/or after school time have showed no negative effect on academic achievement (1,2). In fact, several studies support that regular physical activity might improve cognitive performance and brain functioning, which, in turn, would positively affect academic performance (3–5). Several authors concluded that preadolescent children with better fitness have a better cognition, particularly the part of cognition related to executive control (3–5), but also learning and memory (6). They also observed differences in brain functioning, asmeasured by functional neuroimaging and electrophysiological techniques, and in brain structures, as measured by magnetic resonance imaging (MRI) voxel based morphometry, concluding that fitter children have healthier brains (7–9).

There are several studies looking at the effect of exercise on cognitive performance and academic achievement in children (1,3–5), however, very little is known about how exercise affect brain function and structure in children. To our knowledge, three randomized controlled trials have been conducted on this topic in young people. These studies have used neuroelectric and neuroimaging techniques. Monti et al. (FitKids study) (10), observed that a 9 months intervention, of high intensity aerobic exercise 2h/day x 5 days/week, improved relational memory in preadolescents children, as measured by an eye-movement method. This program also had a positive effect on working memory and task preparation processes as measured by event-related brain potentials (11). This intervention also decreased anterior prefrontal cortex activation from pre-test to post-test, measured by functional magnetic resonance imaging, which reflects a more mature brain function in children who underwent the exercise program (12). Davis et al. (Smart study), observed that a 3 months aerobic exercise program improved executive function, increased bilateral prefrontal cortex activity and reduced bilateral posterior parietal cortex activity (13). The same research group has recently reported the main findings of an 8-month aerobic exercise-based trial (14). The intervention decreased activation in several brain regions on an antisaccade task compared to the control group, possibly reflecting increased efficiency. Such intervention additionally increased activation in both the incongruent versus fixation and incongruent versus congruent contrasts of a flanker task compared to the control group, possibly reflecting greater flexible modulation of cognitive control (14). Finally, this exercise program also positively influenced resting state synchrony, which reflects coherence in the functional organization of the brain independent of task performance (15).

This field is in its infancy and many questions remain to be answered. The ActiveBrains project will use cutting-edge technologies to address novel and sound research questions, such as how exercise influences the volume and functionality of specific brain regions, and networks related with memory and executive function, such us the prefrontal cortex, specially anterior cingulate and dorsolateral, hippocampus, posterior parietal, and basal ganglia.

#### 2.2. State of the art and rationale for the secondary aims

The prevalence of pediatric obesity has reached epidemic proportions in most of developed and developing countries and it is a major public health problem. In Spain, pediatric obesity is of special concern. According to the data from the World Obesity Federation ([www.worldobesity.org](http://www.worldobesity.org)), Spain leads the ranking of overweight/obesity in children aged 7-11 years in Europe, together with Malta and Sicily. Recently, we have objectively assessed physical activity levels in a sample of adolescents from 10 different regions in Europe (9 countries) and observed that adolescents from south Europe (including Spain, Italy and Greece) were less active and more sedentary than adolescents from central-north Europe (16). The low activity levels observed in southern Europe might be responsible, at least in part, of the high prevalence of overweight/obesity and related cardiovascular disorders in southern Europe. Closely linked to obesity an important number of risk factors have been identified in children and increase their future risk for chronic diseases. Insulin resistance and other metabolic alterations in childhood have been consistently linked to paediatric obesity, and further studies are still needed to better understand which type of exercise is more efficient in reducing both adiposity and metabolic risk. In the present study, we will include a complete set of cardio-metabolic risk factors (blood pressure, lipid profile, insulin resistance, etc.), in order to study the effect of the exercise intervention on these factors.

Osteoporosis, and the associated fractures, is another major health problem. The economic burden of osteoporosis in Europe is higher than any kind of cancer (except lung cancer) or chronic cardiorespiratory diseases(17). Although the onset of osteoporosis is an adult disease, early prevention remains the most ef- fective public health action. In this context, it has been consistently demonstrated that acquiring a high bone mass pick during childhood and adolescence, which can be largely influenced by exercise, is a key determinant of adult skeletal health (18). The ActiveBrains project will study the effect of our exercise-based intervention on bone mass and density.

Mental health is a major component of overall health, as defined by the World Health Organization (WHO). A key factor for a healthy mental status is a good management of chronic stress. The current evidence from prospective observational studies, both natural experiments examining real-life stressors and laboratory experiments, shows that stress modifies disease-relevant biological processes in humans. Moreover, experimental studies with animals strongly support a causal link between stress and disease, particularly, depression and cardiovascular disease (19). Several studies conducted in adults showed that exercise can reduce stress levels. However, the nature of stress in adults and in children is different, and little is known about how exercise might affect stress levels in children (20). Observational evidence suggests that high cardiorespiratory fitness in adulthood predicts lower risk of depression, both in cross-sectional and longitudinal studies (21). Likewise, adolescents with lower cardiorespiratory fitness level are more likely to have a diagnosis of psychosis (22) or schizophrenia (23,24).

Taking together the observational evidence on physical and mental health described above (both cross-sectional and longitudinal studies), well-design randomized controlled trials are needed. The ActiveBrains Project will contribute to the understanding of the causal effect of exercise not only on cognition and brain (primary aim), but also on a relevant set of physical and mental health outcomes (secondary aims).

#### 2.3. Capacity of the principal investigator to successfully carry out this project

The Principal Investigator (PI) is a young but already internationally well-known researcher on the field of physical activity, fitness and health in young people, as shown in his CV. Dr. Ortega has participated in major coordinated national and European research projects in the last years (i.e. AVENA, HELENA, EYHS and ALPHA projects). Particularly, the HELENA and ALPHA are EU-funded projects in which the applicant had a major management and scientific role. In the HELENA study, he was the person in charge within the group for the physical fitness assessment in the 10 European cities involved in the project. In the ALPHA study, he was the project secretary, as well as actively involved for the objective measurement of physical activity (through accelerometry), as well as coordinator of the fitness validity sub-project.

Regarding cognition (major outcome of this study), the applicant has recently coordinated a pilot study to determine the effect of exercise on fitness and cognition in adolescents. We observed that doubling the number of physical education sessions per week (i.e. 4 sessions/week) and their intensity had a positive effect on both academic achievement and cognitive performance (25). The findings strongly supported the key role of the intensity of the exercise to achieve improvements in cognition, something that has been taken into account for the exercise program of the present project. The PI has also examined how socioeconomic factors and sleep relates to cognitive performance in adolescents (26,27).

The PI has a large experience on the physical health outcomes included in this project (first set of secondary outcomes). As an example, in a recent longitudinal study, the PI leaded an article showing that improvements in cardiorespiratory fitness across puberty reduced the risk of developing overweight/obesity 6 years later (28). Recently, we reported that higher physical activity level might attenuated the adverse effect of a low birth weight on insulin resistance (29). A collaborative study with the University of South Carolina concluded that there is a subset of obese people who is metabolically healthy and, once cardiorespiratory fitness is taken into account, this subset of obese people has a lower risk of cardiovascular and cancer mortality, compared with the rest of obese people (30). In relation with bone health, the applicant has supervised a PhD Thesis examining the role of physical activity on youth’s bone health. The applicant is a co-author of the largest meta-analysis ever conducted on gene-environment interactions on osteoporosis, as part of the EU-funded project, the GEFOS consortium (GEnetic Factors for OSteoporosis Consortium). The analyses are being conducted on more than 50 different studies from Europe, USA and Australia (pooled N=150,000). The results will have a major impact on the understanding of how environmental factors, such as physical activity, interacts with genetics in relation with bone mass.

Regarding mental health outcomes included in this project (second set of secondary outcomes), the PI has investigated how psychological well-being and cardiorespiratory fitness relates to mortality in US adults (31). Likewise, the applicant has recently leaded a study in collaboration with researchers from Karolinska Institutet in Sweden involving more than 1 million adolescents and observed that adolescents with a very low muscular strength have 15-65% higher risk of having a psychiatric diagnosis (e.g. schizophrenia and mood disorders) and a 20-30% higher risk of premature death (<55 years-old) due to suicide (32).

### **3. AIMS AND HYPOTHESES**

#### 3.1. Overall hypothesis

A 20-week exercise program will have a positive effect on cognition and brain parameters, as well as on a number of physical and mental health outcomes in preadolescent overweight/obese children.

#### 3.2. Overall aim

The general purpose of the present project, namely the ActiveBrains project, is to examine the effects of a 20-week physical exercise program on cognition and brain, as well as on selected physical and mental health outcomes in preadolescent overweight/obese children.

#### 3.3. Specific aims

**PRIMARY AIM**

To examine the effects of a 20-week physical exercise program on brain structure and function, cognitive performance and academic achievement in overweight/obese preadolescent children.

**SECONDARY AIMS**

- Objective 1: To study the effect of this intervention on physical health outcomes: physical fitness, body composition (including bone), glucose and lipid metabolism and blood pressure.
- Objective 2: To study the effect of this intervention on mental health outcomes: perceived and objectively measured stress, and an overall assessment of child behavior and personality, including self-esteem, anxiety and depression.

### **4. METHODOLOGY AND RESEARCH PLAN. TRIAL DESIGN**

#### 4.1. Brief description of facilities and equipment

The University of Granada has all the infrastructures and equipment for a successful development of the ActiveBrains project. The ActiveBrains project aims to study the effect of an exercise program on cognition and brain, as well as on selected physical and mental health outcomes. The work to be developed has 4 main pillars: 1) the exercise intervention itself; 2) assessment of cognitive parameters, as well as neuroelectric and neuroimaging measures; 3) assessment of physical health outcomes and 4) assessment of mental health outcomes.

The PI of this project has gathered a lot of experience on exercise intervention and assessment of physical health outcomes (pillars 1 and 3) from the previous projects in which he has participated (AVENA, HELENA, EYHS, ALPHA, EDUFIT). The University of Granada has now an impressive new facility, namely the Technology Park of Health Sciences of Granada (http://en.ptsgranada.com/). This Park is an area of over 625,000 m² that brings together the infrastructure and quality services to the general objectives which aims: to become a space for teaching excellence, care, research and business, specializing on life sciences. The Park consists of a number of Research Centers and Institutes equipped with the latest advances in technology. One of these Research Centers is the newly built Research Institute of Sports and Health (iMUDS): <https://uceens.ugr.es/sobre-uceens/sedes/instituto-mixto-universitario-deporte-y-salud/>. The constructions for this Institute finished in August 2013. The most advance equipment in different areas of sport sciences has been ordered, received and installed already at the Institute. The setting up of the new equipment will be finished by November 31st, 2013. This means that both infrastructures and equipment will be fully ready and working long before this project would start. The PI, because of his marked research profile, has been selected to settle his office in the Institute, which will happen in December 2013. The exercise program, assessment of physical fitness measures and body composition for the ActiveBrains project will be performed in this Institute using the latest and more advance technology. As an example, maximal exercise test will be conducted in the Institute, using a gas analyzer (General Electric) and the h-p-cosmos treadmill. Likewise, body composition analyses will be conducted using the most known densitometer in the world, the Discovery from Hologic, using pediatric software.

Biochemical analysis (part of pillar 3) will be done in close collaboration with the group headed by Prof. Angel Gil and Dr. Conception Aguilera, and represented in this project by Dr. Carolina Gómez. This research group is settled at the Research Institute of Biomedicine in Granada, which is next to (500m apart) the Research Institute in Sport Sciences, where the main part of the project will be physically carried out. The physical proximity is very convenient and improves efficiency when transporting sample and between-researcher communication (e.g. frequent meetings).

In addition to the Research Institute of Sports and Health and the Research Institute of Biomedicine mentioned above, there is another new center just opened in the University of Granada, the Research Institute of Brain, Mind and Behavior. The assessment of cognition, brain measures and mental health will be carried out in this new Institute (Pillars 2 and 4). This Institute has been equipped with the most recent and up-to-date technologies, including the equipment required for the electroencephalogram and functional magnetic resonance imaging. Remarkably, the magnetic resonance machine just bought is one of the most advance and expensive at the moment (total cost = 1.7 millions of Euros), and will be available to be used in the present project. The center will have expert employees to handle these devices (technicians), and every examination will have an associated cost which is included in the budget of this project.

#### 4.2. Study sample and design

The ActiveBrains project is an individual randomized controlled trial (1:1) that aims to examine the effects of a 20-weeks physical exercise program on brain structure and function, cognitive performance, academic achievement and physical and mental health in overweight/obese children. Participants with overweight/obesity, meeting the eligibility criteria, are included in our study. The ActiveBrains project has been approved by the Review Committee for Research Involving Human Subjects at the University of Granada (Reference: 848, February 2014), and registered in the ClinicalTrials.gov (Identifier: NCT02295072). A total of 110 eligible (see inclusion criteria bellow) participants will be randomized into exercise group vs. control group. For feasibility reasons, the study is conducted in 3 waves: during the first academic year of the project (i.e. 2014–2015) we prepare protocols, set-up measurements techniques, and enroll in the study the first 20 children (50% allocated to each group); at the beginning of the following academic year (i.e. 2015–2016) we enroll another 45 children (50% allocated to each group); and later on that academic year we enroll the remaining 45 children (50% allocated to each group), summing up with these 3 waves the aimed sample size of 110 participants. The control group receives regular physical education sessions (2 per week). Both exercise and control groups receive a pamphlet with recommendations for an active lifestyle and healthy eating habits. To study the extent to which the effect of the intervention remains or disappears once the formal intervention is finished, we will do a third evaluation 8 months after the intervention has finished. In the case that additional budget is obtained and the project prolonged, we will conduct the third evaluation in the whole sample.

The control group will receive the usual physical education sessions (2 per week). Based on existing literature, we believe that exercise is potentially beneficial for the physical and mental health of the overweight/obese children participating, and will not be definitively harmful for them. Therefore, we decided to use the waitlist control group strategy, also used in previous studies in this field (10,11), which will be time and effort consuming, but it will increase moral and ethical values of the project. This strategy implies that the wait-list control group also receives the exercise program but later, after all the assessments of the potential effect of the program have been completed.

4.3. Rationale for the age and weight status group selected for the study

We focus on overweight/obese children aged 8 to 11.9 years. The age group selection was made on the basis that childhood is a critical period where brain function, cognition, obesity and several comorbidities are still under development. Early stimulation on brain structure and functioning might have long term effects. We also selected a preadolescent sample since adolescence and pubertal physiological and psychological changes are dramatic, the speed of changes/maturation differs among individuals, and it is difficult to control these processes and confounding factors. It is known that overweight/obesity is related with a large number of adverse conditions, including metabolic dysfunction, poorer mental health (e.g. high depression risk, lower self-esteem) and recent reports support that excess in adiposity relates to worse cognition (11,33–35).

#### 4.4. Sample size and power

Statistical Package for Social Sciences (IBM SPSS Statistics for Windows, version 20.0; Armonk, NY, USA) was used for calculations. It is difficult to estimate the appropriate sample size and power of our study since there is little information about previous exercise interventions on many of the outcomes here studied. Nevertheless, we have done an estimation based on a previous cross-sectional study and including some assumptions. This cross-sectional study observed a large effect size difference (Cohen-d=0.6) in brain structures (i.e. hippocampus) between children with low vs. high cardiorespiratory level in children (36). For this effect size, an alpha error of 5% and power of 80%, 60 people (30 in each group) would be necessary. However, in this study the differences between the low and high fitness groups in cardiorespiratory fitness (36.4 vs. 51.5 ml/kg/min, respectively) were large (i.e. Cohen-d=3.5) due to the fact that only both fitness extremes were included in the analysis (participants with a middle fitness level were excluded). Considering that an intervention study will lead to a smaller effect size than that between extreme groups reported above in the cross-section study, and also a potential dropout rate of 10% observed in similar studies (37), this project would need 100 participants for a 80% power and an alpha error of 5%. If the economic budget allows it, we will try to recruit 10 more participants to a maximum of 110. This sample size is feasible and realistic based on our previous experiences involving electroencephalograms and magnetic resonance imaging.

#### 4.5. Inclusion/exclusion criteria

The eligibility criteria to participate in this study are: 1) children aged 8 to 11.9 years; 2) in the case of girls, pre-menstrual at the moment of baseline assessments; 3) classified as overweight or obese at baseline based on sex and age specific World Obesity Federation cut-off points (38,39); 4) not to have any physical disability or neurological disorder that impeded exercise; 5) not to use medications that influenced central nervous system function; 6) right-handed as assessed by the Edinburgh inventory (40); and 7) no previous diagnosis of attention-deficit hyperactivity disorder (ADHD) and a score above the 85^th^ percentile as measured by the ADHD rating scale (41). Children with psychiatric diagnosis at baseline or during the trial will also be excluded from the analyses. Every child randomized to the exercise group will go through complete medical examination, and children with any medical condition that would affect the results of the evaluations or that limit the normal capacity to do exercise will be excluded.

#### 4.6. Recruitment and randomization

The recruitment process will start by contacting families of children with overweight/obesity from databases at the Unit of Pediatrics of the University Hospitals San Cecilio and Virgen de las Nieves (Granada, Spain). Additional strategies include contacting the head teacher of both, public and private schools of Granada to spread informative pamphlets. Furthermore, advertising related to the project will be broadcasted in the local media through newspaper, radio, and television outlets. The current Spanish healthy system cannot afford a free of charge exercise program for all those overweight/obese children who would benefit from it. Therefore, the ActiveBrains project is offering to overweight/obese children and their families a well-designed and controlled exercise program that is a recommended “treatment” by paediatricians without any cost for the families. Because of the wait-list control group strategy, all the participants benefit from the program. Three paediatricians and two nurses are involved in the study and they do a basic clinical examination, including physical examination, weight, height, blood pressure and pubertal status assessment.

Simple random allocation of participants into exercise or control groups will be performed with a ration 1:1 using a computer random number generator in Statistical Package for Social Sciences (IBM SPSS Statistics for Windows, version 20.0; Armonk, NY, USA) by a “blinded” individual not involved in the exercise sessions nor outcome evaluations (FBO). This method allows for the equal probability of being allocated to one group or another. To reduce the risk of bias, several protocols will be followed: 1) the computer random generation will be conducted by a person not involved in the outcome evaluations; 2) randomization will be performed immediately after the baseline evaluation; and 3) the physical trainers running the exercise program will not be involved in the outcome evaluations or randomization. Randomization is done immediately after the baseline assessment is completed in order to reduce the risk of bias during the assessment.

#### 4.7. Intervention

The intervention consists of 20-weeks exercise program. It is our aim to test whether the internationally accepted physical activity guidelines (<http://www.health.gov/paguidelines/>) are effective to improve brain structure and function and cognition as well as physical and mental health in overweight/obese children. The guidelines recommend children to exercise daily, and, therefore, we offer the possibility to attend to the exercise program daily from Monday to Friday (i.e. 5 sessions/week, 90 min/session). However, we are aware that in addition to our exercise program children in Spain usually have 2 physical education sessions per week and many of them also have sport-based after-school activities twice per week. Taking this into account, we will inform parents and participants that our exercise program is offered 5 sessions per week, but the minimum attendance recommended is 3 times per week. The physical exercise program will be conducted on a group basis (i.e., 3 waves of intervention) and based on active games, with a noticeable emphasis on the playful component in order to increase adherence to the program. Each session is structured in four parts: 1) a 5-10 min warm-up consisting of 1-2 physical games of 5 min each; 2) a 60-min aerobic part consisting of around four to five physical multi-games requiring moderate-to-vigorous intensities, with special emphasis on high-intensity activities; 3) a 20-min resistance training part consisting of muscle- and bone-strengthening game-based activities. The resistance part includes exercises involving large-muscle-groups for which therabands, fitballs as well as participant’s own bodyweight were used; and 4) a 5-10 min cool-down part consisting of stretching and relaxation exercises.

The intensity of the exercise program is monitored in all children across the whole exercise program. Participants’ progress relative to exercise intensity will be checked weekly by trained personnel to: (i) adapt the intensity of the program progressively according to the improvements of the participants; and (ii) to identify whether any child is training at lower intensities than the rest of the group, thus requiring higher motivation during the exercise sessions. The heart rate data will be recorded during both the aerobic and the resistance training components. Every child will wear the same HR monitor (POLAR RS300X, Polar Electro Oy Inc., Kempele, Finland) individually programmed based on their maximum heart rate previously achieved in the maximal incremental test (see test measurement description below). Moreover, we will also program the monitors individually at 80% of the maximal heart rate and at the level of the anaerobic threshold, so that we can later obtain the accumulated time over the 80% of the maximum HR and over the anaerobic threshold.

#### 4.8. Rationale for the exercise program design

Previous literature showed that aerobic exercise is the most effective type of exercise to improve brain and cognition (42); consequently, this project has a special emphasis on aerobic exercise. However, since this project is an intervention aiming to achieve a benefit not only on children’s brain and cognition, but also on general physical and mental health, we also include activities that enhance muscular strength and speed-agility, as well as activities that strengthen children’s bones. Evidence, mainly based on observational studies, supports a link between aerobic fitness and cognition, but there is little information for other components of physical fitness, i.e. muscular strength or speed-agility. One observational study by Castelli et al. reported an association between cardiorespiratory fitness and academic achievement, but not between muscular strength and academic achievement in children (42).

High intensity exercise is an important goal to achieve in every session of the program since there are reasons to believe that this kind of exercise is the most effective for different health outcomes, and it seems also for cognition and brain. As an example, Castelli et al., observed that preadolescents who spent more time of the sessions at high intensities (above the 80% of heart rate maximum) showed larger improvements in cognitive performance (43). In order to obtain objective and high quality data on the relative intensity and physiological demands of our physical exercise program every child in the exercise group wear a heart rate monitor during all the sessions of the program.

The length of the intervention (20-weeks during a school-academic year) is within the timeframe used in the previous randomized controlled trials on this topic, i.e. 3 months (13) and 9 months (10). It has often been reported that when an intervention program is implemented, it might have a compensatory effect, so that the participants stop doing other physical activities that they would have done otherwise. To study this issue, we will assess physical activity using activity monitors (i.e. accelerometers) over 7 days at 3 different time-points during the study: baseline, middle of the intervention and post- intervention.

#### 4.9. Strategies to enhance compliance and adherence to the program

Participants and their parents will be verbally motivated to participate in the program and to attend to all the assessment and exercise program sessions. Our goal was that children attend at least 3 sessions per week. However, we encourage the children and their families to attend 4 or 5 sessions per week whenever possible. Children who complete successfully the program get a “certificate” as “successful completers”. Children are the key part of this project and they deserve acknowledgements from this project and for their positive attitude and willingness (and their family) to participate in this project.

#### 4.10. Primary outcomes

The full set of primary and secondary outcomes are assessed twice, immediately before and after the 20-weeks exercise program. A third assessment will be conducted 8 months after the intervention finishes.

1. **FUNCTIONAL AND STRUCTURAL MAGNETIC RESONANCE IMAGING**

The neuroimaging techniques, particularly structural and functional MRI, provide a great opportunity to deepen into the field of exercise and brain structure and function. In the present project, we plan to conduct the following protocol using structural and functional 3.0 Tesla Siemens Magnetom Tim Trio scanner MRI (Siemens Medical Solutions, Erlangen, Germany) with a 32-channel head coil (total time = 40–45 min, including rest between MRI sequences):

a. High resolution scanning (7min 31 s). This provides structural information of the whole brain. We will look at individual changes in the whole brain since much needs still to be discovered about the effect of exercise on brain. Based on existing literature (5,9,36), we also specifically analyze the effect of the program on the prefrontal cortex, anterior cingulate cortex, dorsolateral prefrontal cortex, basal ganglia (dorsal and ventral striatum; and the region globus pallidus), insular and parietal cortices, superior frontal sulcus and hippocampus (special interest on the dentate gyrus region). The hippocampus, which is a subcortical brain region that is vital for relational memory, demonstrates both a high plasticity and a high capacity for synaptic modulation. High-resolution, T1-weighted images will be acquired using a 3D MPRAGE (magnetization-prepared rapid gradient-echo) protocol. The acquisition parameters are the following: repetition time (TR) = 2,300 ms; echo time (TE) = 3.1 ms; inversion time (TI) = 900 ms; flip angle = 9º; field of view (FOV) = 256 x 256; acquisition matrix = 320 x 320, 208 slices; resolution = 0.8 x 0.8 x 0.8 mm; and scan duration = 6 min and 34s (44,45).
b. Functional magnetic resonance in resting state (5 min 26 s). This provides information about the effect of the program on brain functioning in a resting situation. The resting-state functional MRI data consists of a series of 160 scans acquired using a Gradient Echo Pulse Sequence while participants rest with eyes closed. The parameters are as follows: TR = 2000 ms, TE = 25 ms, flip angle = 80º, FOV = 240 mm, acquisition matrix= 240 x 240, 35 slices, resolution = 3.5 x 3.5 x 3.5 mm, and scan duration of 5 min and 26s.
c. Diffusion Tensor Imaging (DTI) (5 min 18 s). This provides information about neuronal connectivity and fibre structure in white matter.

2. **NEUROELECTRIC MEASUREMENTS**
   a. We use the modified flanker task (MFT) to assess inhibition and delayed nonmatch to sample (DNMS) task to evaluate learning and memory. Simultaneously with these tasks, we measure selected components of the event-related brain potentials. For example, we analyze the amplitude and latency of P3 (also called P300). Higher P3 amplitude is considered an indicator of a better ability to recruit attentional resources (46), while a lower P3 latency is considered an indicator of faster cognitive processing speed (46).
   b. Event-related brain potentials, obtained from the electroencephalogram, assess aspects of human information processing, and have provided insight into the underlying brain mechanisms involved in cognitive function beyond that of overt behavioral task performance. In our study, we use the ActiveTwo System of Biosemi (64-channel, DC amplifier, 24-bit resolution, biopotential measure- ment systemwith Active Electrodes). The complete assessment (i.e. preparation plus the time devoted to the tasks) takes about 80–90 min.
3. **COGNITIVE PERFORMANCE**

We focus on a set of tests, which are internationally well-known and validated in children, mainly aiming to assess executive function (also called cognitive control or executive control), which has shown to be the most sensitive to be influenced by exercise (3–5). The specific constructs to be measured and tests chosen for such purpose are (total testing time 60 min approximately):

a. K-BIT, Kaufman Brief Intelligence test that measures both verbal and non-verbal intelligence (47).
b. The Design Fluency Test (48–50) measures one's initiation of problem-solving behavior, fluency in generating visual patterns, creativity in drawing new designs, simultaneous processing in drawing the designs while observing the rules and restrictions of the task, and inhibiting previously drawn responses. This test belongs to the test battery DKEFS, Delis-Kaplan Executive Functions System.
c. The Stroop Color-Word Test is used to measure cognitive inhibition (DKEFS) (51).
d. The Zoo Map Test measures spatial planning, rule learning, inhibition of impulsive and perseverative responding, and the ability to establish and maintain instructional set.
e. The Trail Making Test measures flexibility of thinking on a visual-motor sequencing task (DKEFS) (52).
4. **ACADEMIC ACHIEVEMENT**

We evaluate this important dimension using two methods:

a. First, we use the final school grades obtained the year before the intervention (e.g. June 2014) and after the intervention (e.g. June 2015). This is the final result of many factors and what parents and society worry about the most. However, there is a potential systematic bias among different schools and teachers, so it is optimal to complement this measure with a standardized measure of academic achievement (see below).
b. Second, we additionally use the Batería III Woodcock-Muñoz™ (age range 2 to 90 years-old), which is the Spanish adaptation/translation of the Woodcock-Johnson III®(WJ III®) (53). This battery has a specific academic achievement battery (testing time: 70–90 min) that consist of the following tests: 3 tests of reading, 2 tests of oral language, 3 tests of mathematics, 3 tests of written language and 1 test of academic knowledge (i.e. Science, Social Science and Humanities).
5. **OTHER BRAIN-RELATED MEASURES**

There is increasing evidence supporting that beneficial effects of exercise on the body and brain are mediated by the brain-derived neurotrophic factor (BDNF) (54). Within this context, findings from recent reviews and meta-analysis support the role of exercise as a strategy to increase the BDNF activity in humans (55,56). The BDNF acts on certain neurons of the central nervous system and the peripheral nervous system, helping to support the survival of existing neurons and encourage the growth and differentiation of new neurons and synapses. The BDNF has shown to act together with another important protein, the Epidermal Growth Factor (EGF), activating Neuronal m-Calpain, which plays a significant role in synaptic plasticity, cell motility, and neurodegeneration. The circulating blood BDNF is measured in serum using the RayBio Human BDNF ELISA (Enzyme- Linked Immunosorbent Assay) kit.

#### 4.11. Secondary outcomes

1. **PHYSICAL HEALTH OUTCOMES**
a. Physical fitness (cardiorespiratory fitness, muscular strength and speed-agility) is assessed following the ALPHA fitness test battery (57). Briefly, cardiorespiratory fitness is assessed by the 20m shuttle run test; muscular strength is assessed by handgrip strength test and standing long jump test and speed-agility is assessed by 4 × 10m shuttle run test. Cardiorespiratory fitness is additionally assessed using a gas analyser (General Electric Corporation) while performing and maximal incremental treadmill (hp-cosmos ergometer) test modified for poorly fit children (37). Participants walk on a treadmill at a constant speed (4.8 km/h) with a 6% slope with grade increments of 1% every minute until volitional exhaustion. Children are encouraged to walk as long as they can. This test is done by Sport Medical Doctors from the Andalusian Centre of Sport Medicine. Maximal oxygen consumption (VO_2_max, ml/kg/min), heart rate and respiratory exchange ratio (RER) are recorded each 30 second. Ratings of perceived exertion (RPE) using children's OMNI scale (58) is registered at the end of each 1 minute stage. VO_2_max is confirmed when meeting the 3 out of 4 following criteria: volitional fatigue (N8 points in the OMNI scale), a plateau in VO_2_max during the last two exercise work rates (2.0 mL·kg−1·min−1), achieving >85% of age-predicted heart rate maximum (59) and a RER of ≥1.10. In addition to the field-based tests from the ALPHA battery, we assess muscular strength in laboratory condition using pneumatic resistance machines (Keiser Sports Health, Fresno, CA, USA). According to previous studies in paediatric population, we determine each participant 1 repetition maximum (1RM) strength in bench press and leg press tests (60–63). The 1RM is recorded as the maximum resistance that is able to lift throughout the full range of motion. Participants receive familiarization sessions before testing session in order to ensure an adequate technique (i.e. controlled movements and proper breathing). Using standardized procedures (63), before attempting a 1RM, participants perform 6 repetitions with a light load and 3 repetitions with a heavier load (50–90% estimated 1 RM). Then, a series of single repetitions with increasing loads (0.5–2.3 kg for bench press and 10–20 kg for leg press) are performed. The 1 RM is determined when participants falling short of the full range of motion on at least 2 not consecutive attempts. During testing, participants have 3–5 minutes of resting between trials. RPE using children's OMNI-Resistance Exercise scale (64) is registered for each attempt. Moreover, during all testing procedures the examiner asks “How do you feel?”, “Is the load light, medium or heavy?” and “Could you lift more?” to aid the progression of the 1 RM trials. 1 RM of bench press and leg press perform in the 80% of the whole sample.
b. Subjective perception of the level of physical fitness is measured by International Fitness Scale (IFIS). The IFIS is a reliable, simple and short self-administered scale composed of five Likert-scale questions about the perceived youth overall fitness and the main components: cardiorespiratory fitness, muscular fitness, speed-agility, and flexibility in comparison with their friends’ physical fitness (very poor, poor, average, good and very good) (65). IFIS has shown a good validity against measured fitness in children of this age range (66).
c. Physical activity and sedentary time are assessed by accelerometers. A tri-axial accelerometer (Actigraph GT3x, Pensacola, FL, USA) is used to assess physical activity and sedentary time over 7 consecutive days. Participants are instructed to wear two devices: one attached using an elastic band to the right hip and one attached to the non-dominant wrist (which in all cases is the left one). Children wear the accelerometers 24 h and remove it only while bathing or swimming. Also, children have a log in order to record the time when they go to bed, wake up, and remove the device.
d. Self-report of physical activity levels and sedentary behaviors in youth are assessed by the Youth Activity Profile — Spain (YAP-S). The YAP-S was designed to be a self-administered 7-day recall questionnaire suitable for use in children (67). Under the umbrella of the ActiveBrains project, we translated (into Spanish) and back-translated (to test possible deviations from the English original version), as well as culturally adapted, in collaboration with the original authors of the YAP. Calibration studies on the YAP-S is being conducted during our study using the accelerometer data collected the week immediately before. The test–retest reliability of this tool is also being tested in a different sample. The test includes 15 items that ask about activity at school, activity out of school, and sedentary habits.
e. Commuting to and from school is evaluated by a self-reported questionnaire. Participants answer the following questions “How do you usually travel to school?” and “How do you usually go back to school?”. Also, the second set of questions refers to the way of commuting to and from school during a week. The responses can be: by walk, bike, motorbike, car, bus, several transports or other transport requesting in these cases.
f. Anthropometrics measurements. Weight, height, body mass index, waist circumference and triceps and subscapular skinfolds thickness are evaluated. Body weight is measured with an electronic scale (SECA 861, Hamburg, Germany) and height (cm) with a stadiometer (SECA 225, Hamburg, Germany). In addition, we assess peak height velocity (PHV) as an accurate and discriminant measure of maturational status (68). PHV was calculated from age and anthropometric variables following Moore’s equations (69). Years from PHV were calculated by subtracting the age of PHV from chronological age, so that it is interpreted as how many years from the PHV offset a person is, with a value ranging from negative values (before the PHV; less mature) to positive values (after the PHV; more mature).
g. Body composition and bone mineral density are assessed by Dual- energy X-ray absorptiometry (DXA, Discovery densitometer from Hologic), following protocols used in our previous studies (70,71). Likewise, fat mass, fat-free mass, total body water and bone mass are assessed by the TANITA bioimpedance balance (BC-418 MA, TANITA International Division, TANITA, UK).
h. Traditional cardio-metabolic risk factors include a complete set of risk factors as markers of lipid profile (triglycerides and total-, HDL and LDL-cholesterol), blood pressure (following standard procedures), and insulin resistance (glucose and insulin, homeostasis model assessment, HOMA). Blood sample (in fasting condition) is collected before and after the exercise program and 8 months after the intervention finishes. It is divided in 4 tubes in order to obtain plasma aliquots, haematological sample, serum aliquots and biochemical parameters.
i. DNA (Deoxyribonucleic acid) and RNA (Ribonucleic acid) analyses. We freeze the blood, plasma, serum samples, so that they can be analyzed later when the intervention and outcome assessments are finished. This strategy provides us with the most up- to-date list of candidate genes for the different study outcomes.
2. **MENTAL HEALTH OUTCOMES**
a. Stress will be measured by the Children's Daily Stress Inventory (CDSI), a measure that assesses daily stress in primary school children. The inventory was validated in a sample of 1094 primary school Spanish students (72). The final version includes 22 dichotomous items covering the areas of health, school/peers, and family. Also, parents have to complete this questionnaire thinking as if they were their own children. An objective measure of stress is heart rate variability, which is defined as the variations of both instantaneous heart rate and RR intervals on the electrocardiogram. In a healthy situation, considerable variability (i.e. within the range) reflects the heart's ability to adequately respond to physiological and environmental stimuli. There is accumulating evidence indicating that a lowheart rate variability is an indicator of chronic stress (73). Each child is individually examined in a quiet room in the supine position for 10 min and comforting music is played to encourage a relaxation status. We use the top-class heart rate monitors, Polar RS800CX (Kempele, Finland), for the measurement (i.e. 10 min) of heart rate variability parameters. This device has established validity compared to the gold standard of an electrocardiogram device in children (74).
b. Childhood trait anxiety is measured with the trait score of the State–Trait Anxiety Inventory for Children (STAIC-T). It is a 20 item self- administered instrument which is widely used, reliable (Cronbach alpha=0.94) and extensively validated (75). Parents complete the same questionnaire according to their own perceptions about their children.
c. Children Depression Inventory (CDI) is used to assess depression. The test has five scales (negative mood, ineffectiveness, anhedonia, negative self-esteem and interpersonal problems) which are based on the children experiences (76). It comprises 27 items. Parents complete the same questionnaire according to their own perceptions about their children.
d. Self-concept is evaluated with the Self-concept form 5 (AF5). Physical, labor, social, family and emotional dimensions are assessed with this test. Children have to complete 30 items with a response scale between 1 and 10. Psychometrics properties showed an internal consistency for the AF5 of 0.83 for all dimensions, while for the individual dimensions were 0.90 for academic, 0.69 for social, 0.71 for emotional, 0.78 for family and 0.77 for physical (77).
e. Body image is evaluated by the Children's Body Image Scale (CBIS) for preadolescent children (78). The test use gender specific figures posed in the anatomical position. Each figure is a modified photo- graph of an anonymized, pre-pubescent boy or girl with a BMI within the specified range for one of seven percentiles (3rd, 10th, 25th, 50th, 75th, 85th, 97th) or body composition categories established by the WHO (formerly named the International Obesity Task Force) cut-off (79). Participants have to select their perceived and ideal body size from 7 figures representing body sizes from underweight to obesity. Body dissatisfaction is obtained by subtracting the ratings of participants’ ideal body size from their perceived current body size.
f. Self-efficacy is assessed using the General Self-Efficacy Scale (GSE). The GSE is a 10 items scale designed to evaluate the positive belief in one's ability to achieve goals or deal with the challenges across various stressful situations. Reliability analyses showed that Cronbach alpha ranged from0.76 to 0.90 (80).
g. Self-esteem is assessed by the Rosenberg Self-Esteem scale (81), which is composed by 10 items. Participants have to mark if they agree or disagree with each statement. The Rosenberg scale is widely used in studies with children and adolescents (82,83). To be more sensitive in specific parts of mental health, we decided to assess self- concept and self-esteem separately.
h. The Positive Affect Schedule for Children (PANAS-C) evaluates in 20 items positive affect and negative affect in children (84). The PANAS- C has shown a Cronbach alpha from 0.87 to 0.90 for the positive affect subscale and from 0.87 to 0.94 for the negative affect subscale (85). Parents complete the same questionnaire according to their own perceptions about their children.
i. Happiness is measured by the Subjective Happiness Scale (SHS) (86), which includes 4 items. The Spanish version of SHS showed an adequate internal consistency, appropriate test–retest reliability and convergent validity (87).
j. Dispositional optimism is evaluated with the Life Orientation Test—Revised (LOT-R) (88). The test comprises 10 statements. LOT- R is a useful, valid and reliable self-report measure to properly assess optimism in adolescent (89).
k. As an overall measure of behaviour and personality traits of the children, we use the Behavioural Assessment System for Children (BASC), the version reported by parents, which measures relevant dimensions such as Aggression, Hyperactivity, Conduct Problems, Anxiety, Depression, Somatization, Attention Problems, Atypicality, Withdrawal, Adaptability, Leadership and Social Skills.
3. **OTHER OUTCOMES**
a. Children attitude and self-perceptions toward physical activity are evaluated with Children's Self-Perceptions of Adequacy in and Predilection for Physical Activity (CSAPPA) questionnaire. The scale has 20 items (7 for adequacy, 10 for predilection and 3 for enjoyment). CSAPPA have shown to be a useful, reliable and valid tool to measure perception of subjects' adequacy in, predilection for, and enjoyment of physical activity in Spanish school context (90).
b. Health-related quality of life is measured by a new, valid and feasible tool called Child Health Utility 9D (CHU9D) (91,92). This test was designed specifically for young people (7–11 years). The CHU9D consists of 9 dimensions: worried, sad, pain, tired, annoyed, school- work/homework, sleep, daily routine, and ability to join in activities with 5 different levels representing increasing levels of severity within each dimension. In addition, CHU9D scores are used in cost-utility analyses (93).
c. Ad-hoc injury questionnaire is applied when the participant present an injury. Questions such as type of injury, description and localization of the injury, modifications of the session due to the injury, grade of the injury, type of exercise in the moment of the injury and how long have been the participant performing the session are registered by the monitor of the program.
d. Sleep quality is assessed by accelerometry (Actigraph GT3x, Pen- sacola, FL, USA). The accelerometer data procedures have been explained above (i.e. physical activity and sedentary section). Additionally, we use the Paediatric Sleep Questionnaire (PSQ) as a measure of night time and sleep behavior, daytime behavior and other possible problems and inattention and hyperactivity. The Spanish version of the PSQ showed internal consistency and high reliability (94). This questionnaire is completed by the parents, who are asked to rate each item according to their child's usual sleep habits.
e. Dietary assessment: two non-consecutive 24-hour recalls are applied referring to weekdays whereby all the foods and drinks consumed on the previous day are recorded to the interviewer. The 24-hour recalls are conducted in presence of the child's parents because they can report dietary intake with more reliability than the children (95) and it has been considered to be the best method to evaluate energy intake in children (from 4 to 11 years) (96). Nutritionists use a photographic manual of food portion size to improve the estimated amount of dietary intake. All the data is registered by the EasyDiet software (© Biocentury, S.L.U. 2016), which is the software supported by the Spanish Association of Dietetics and Nutritionists. Moreover, frequency of food intake is evaluated by food frequency questionnaire (97). It contains questions regarding the average frequency and amounts of 72 selected foods. A nutritionist interviews the child and at least one of parents. The adherence to the Mediterranean diet (AMD) is assessed using the KIDMED questionnaire (98).
f. Perinatal data such as birth weight, birth length and birth head circumference are asked by an ad-hoc questionnaire. Type of breastfeeding and duration and several questions about the mother pregnancy such as weight gain and smoke are recorded by a self-report questionnaire.
g. Cost-effectiveness is assessed by ad-hoc questionnaire. Number of paediatrician’s visits, days in the hospital, medicines intake and its cost and whether their child had had day lost study due to overweight/obese or other health problems during the last 20-weeks are answered by parents.
h. Educational level, profession and socioeconomic status (determined by Family Affluence Scale questionnaire) are asked to parents (99).

#### 4.12. Data analysis plan

Two analysis will be defined: the per-protocol analysis and the intention-to-treat (ITT). We will use the per-protocol principle to report the main findings for all behavioral and MRI outcomes in all children with overweight/obesity that followed these criteria: (1) completed the study and the pre- and post-intervention assessments, and (2) attended at least 70% of the recommended 3 sessions/week (i.e., exercise group). Main analyses will be performed using the per-protocol criteria for two reasons: 1) we are interested in knowing the efficacy rather than effectiveness of our intervention, i.e., we want to know the effects on brain health outcomes when a child actually does the planned exercise program (operationally defined as attending a minimum of 70% of the sessions); and 2) in the field of neuroimaging, with analyses conducted directly on images, it is rarely done and technically difficult, to apply imputation methods on images missing at post-exercise evaluations. Therefore, participants who complete both pre- and post-intervention evaluations are usually included in analyses. Moreover, we will additionally analyze the data using the ITT. Under the ITT principle, we will use multiple imputation for observations lost at post-intervention (100).

The main effects of the exercise program versus control on the study outcomes will be examined taking a previous major RCT that also tested the effect on cognitive outcomes (101) as reference and using ANalysis of COVariance (ANCOVA) including post-intervention outcomes as dependent variables, group (i.e., exercise vs. control) as a fixed factor, and baseline data of the study outcome as covariate. This model indicates the time x group interaction intended to know the effects of the intervention by including the study outcome baseline value as covariate and the post-intervention outcome as dependent. The z-scores for each outcome at post-exercise program will be formed by dividing the difference of the raw score of each participant from the baseline mean by the baseline standard deviation (i.e., (post-exercise individual value – baseline mean) / baseline SD).

MRI data need to be handled using methods specifically developed for this field. Our group has expertise in MRI data analysis and will choose the best approach to every single research question addressed.

The statistical procedures will be performed using the SPSS software (version 20.0, IBM Corporation) and R software (v. 3.1.2, <https://www.cran.r-project.org/>). A significant difference level of P < 0.05 will be set. Additionally, we will investigate which of the significant findings persist after adjustment for multiple testing on the primary outcomes (102).

### **5. CONTINGENCY AND RISK MANAGEMENT PLANS**

There are three main contingency and risk plans to take into account:

1. The study of the effect of exercise on cognition and brain is the most novel, original and scientifically sound part of the present project. However, very little is known about how the brain works and how brain might react to the chronic stimulus of exercise. This part is therefore high impact-high risk. For this reason, the ActiveBrains project has developed a contingency and risk management plan. In addition to the primary aim, the project has been designed to address two other interesting and relevant research questions (secondary aims), particularly how exercise influences selected physical and mental health outcomes. These research questions are also novel; however, there is more previous literature on them which necessary reduces the risk.
2. We are aware that the recruitment of participants for the first wave is challenging, since the study has just started. If for example, 15 instead of 20 participants are recruited by the time the intervention should start, we will go ahead with the intervention and for the second wave 50 children will be recruited instead of 45 planned. Recruitment for the second and third wave is easier, since we have nearly 1 year for recruitment. If number of participants for the first wave is too low, we will use other strategies to increase the recruitment (e.g. visits schools, advertisement in newspapers and websites).
3. Dropouts and adherence during the study. A number of dropouts are expected in any intervention or follow-up study. We have had this into account and the study is powered for 10% dropout rate. Nevertheless, in order to reduce participants’ drop-out and to maintain adherence to the training program, several strategies will be used such as certificates, flexibility in the exercise program which will take into account private commitments, vacations, etc. small and training groups, personalized training and frequent face to face contact.

### **6. FEASIBILITY AND CREDIBILITY OF THE PROJECT**

The feasibility and credibility of the project is warranted due to several factors:

1. The PI has proved over the last 11 years to be a very productive scientist with a large experience and capacity on exercise, physical fitness, obesity and metabolic disorders in children.
2. Additionally the PI has previously worked on cognitive performance (observational and intervention studies) and mental health outcomes, which all together cover most of the outcomes included in the present study.
3. Experimental psychologists will be involved in this project. They have a large experience and knowledge on epidemiological and intervention studies focused on executive function (a major cognitive dimension in this project) and neuroelectric and neuroimaging measures of brain function.
4. Infrastructure/facilities needed for this study are available and ready to be used. As described above, three Research Institutes newly developed and equipped will be involved in this project.
5. The applicant and collaborators have experience in assessing a large number of subjects in epidemiological studies. The sample seems reasonable based on our experience.

### **7. SCIENTIFIC IMPACT OF THE STUDY**

#### 7.1. Scientific impact at a national and international level

Brain is probably one of the most complex and unknown organs in human body. The understanding of the functioning and complexity of the brain is considered by many as one of the main challenges for the 21st century. Previous research was mainly done in animals, due to ethical and methodological issues. However, the latest advances in neuroelectric and neuroimaging technologies will lead to a new era of studies and knowledge. The project aims to be in the front line of the knowledge regarding cognition and brain. Exercise is an easily available and cheap medicine with multiple benefits, but little is still known about its effects on brain. The ActiveBrains project will explore the extent to what exercise is able to improve cognition and brain, at the same time that physical and mental outcomes. The combination of the selected primary and secondary outcomes included in this project will provide new insights on the multidimensional benefits of exercise on youth’s health.

When evaluating the ActiveBrains project, it is important to keep in mind, that knowledge is dynamic, a particularly knowledge on human mind will be developed at a high speed since much is being invested on this topic internationally. A strength of the current project is that we will use one of the most advance magnetic resonance imaging device and will obtain top-quality raw data. When the time for analysis comes, data will be interpreted using the level of understanding about the brain available at that moment. This project will be early in this arena, and data can be re-analyzed in the future when significant advancement are made in the understanding of brain and its functioning.

#### 7.2. Impact on society

Mental performance and the increase of mental illness in children are of growing concern all around the world. In Europe, dysfunctional cognition/cognitive development (perception, memory, intelligence etc.), anxiety, attention deficit hyperactivity disorder, stress, depression and other mental disorders are estimated to affect around 35% of children, resulting in a reduced quality of life and additionally, significant cost impact on European Society through increased treatment and childcare costs. The current project would have impact on European society by providing novel information on the impact of exercise on brain. Ultimately, improved neuro-cognitive function and mental performance of the EU community should reduce the risk of common mental disorders and should increase education, productivity and economic growth.

We expect this project to have a high impact on societies, as previous research activities leaded by the PI have had. As an example, one of the first manuscripts published by the Dr. Ortega on fitness and cardiovascular health received a high attention from the Spanish media and got 3 research awards by the Spanish Society of Cardiology. Similarly, Dr. Ortega recently received the most prestigious award of the University of Granada to the scientific trajectory of young scientists, which is granted by the Social Council of the University illustrating relevance and impact of his research activity on society. Another example is his paper published as first author in the prestigious European Heart Journal. The manuscript got a massive media coverage and was published in more than 300 websites (BBC, CNN, etc.).

### **8. DISSEMATION AND TRANSFERENCE OF FINDINGS**

#### 8.1. Dissemination plan

1. A major pillar of the dissemination plan is to keep a high scientific production in terms of high quality publications. The PI of this project has published more than 190 scientific articles in the last years, being most of them published on top Journals of the field. To highlight is the 3 papers as first author in Journals with impact factor >10. This warrant that the funds invested on this project will be transform into scientific pieces that will reach all around the world.
2. Participation in major International meetings to present the project results.
3. In addition, we will organize one international scientific symposiums in Granada on exercise and cognition/brain in youth during the 2 years that last this project. The symposium will be organized close to the end of the project, to have an overall discussion about the project achievements, as well as to get an update on other related research projects going on. It is important to share and exchange opinions with other top researchers in the field to achieve the research excellence. The University of Granada, the Regional Council for Research and some other entities in Spain support the organization of scientific events.
4. Another fresh and continuous source of dissemination of findings and news related with this project will be the design of a web-site for the project.
5. The University of Granada has a specific service for dissemination of research activities (<https://canal.ugr.es/ugrdivulga/>) that will be very useful to spread out news about relevant findings of this project. In this context, to mention that this applicant is use to deal with media and journalists, both through writing articles for understanding of laydown people and radio-interviews (Spanish and English speaking). As commented above, the manuscript published in the European Heart Journal has received more than 300 news on the online and printed newspapers/magazine (BBC, CNN, The Independent, etc; click on this link as an example: http://www.bbc.co.uk/news/health-19474239), plus a large number of phone interviews from all around the world.

#### 8.2. Transference plan

The project team is planning to do three main transference activities to ensure that most relevant scientific material derived from this project reach the society, particularly non-specialists who do not often access to research information sources (e.g. Pubmed, Web of Sciences). They will be the target for these activities.

1. A first (and almost compulsory) step for increase the visibility of this project as well the transfer of knowledge to the society is the designing of a website. Some people believe that “What is not on the Net, does not exist”. The web domain will potentially be: <http://profith.ugr.es/activebrains?lang=en>; and as a principal page it will have a section of “Latest news”. We expect to use the web also as an informative tool. Overweight/obese children and/or their family might get to know about the project via the website. So, the web will provide with relevant information about the project and contact details to those families who are interested to participate in the project. In separate pages, the web will provide relevant information about the overall background and aim of the project, researcher involved, updated list of scientific publications, links with other related websites or projects.
2. As part of this project we plan to write an article for Wikipedia about exercise and cognition/brain in children (<http://en.wikipedia.org/wiki/Wikipedia:Contributing_to_Wikipedia>). As indicated in Wikipedia website “Since its creation in 2001, Wikipedia has grown rapidly into one of the largest reference websites, attracting 470 million unique visitors monthly as of February 2012. There are more than 77,000 active contributors working on over 22,000,000 articles in 285 languages. As of today, there are 4,055,436 articles in English. Every day, hundreds of thousands of visitors from around the world collectively make tens of thousands of edits and create thousands of new articles to augment the knowledge held by the Wikipedia encyclopedia.” Without a shadow of a doubt, Wikipedia is at the moment one of the most powerful and reliable tools to share knowledge with society. We will make use of it as a major outreach activity.
3. Another powerful Internet Server is Youtube. We have previously used this multimedia online tool for scientific purposes. One of the main outcomes of the ALPHA project, a EU-funded study, was the evidence-based ALPHA fitness test battery available of the website of the project ([www.thealphaproject.net](http://www.thealphaproject.net)). One of the project deliverable was to create a DVD showing how to assess the different physical fitness tests in children and adolescents. Instead of burning DVDs and charge the interested people for them, we uploaded these videos on Youtube, so that everybody in the world has a quick and free access to them (see an example of these videos: <http://www.youtube.com/watch?v=6cVUGoSbHFU&feature=g-hist>). Likewise, we will use Youtube for uploading any visual information that needs to be shared broadly with the public, such as commentaries on articles derived from the projects interviews.
