## Supplement 3 eTables and eFigures for "Effects of the ActiveBrains trial on cardiometabolic and mental health in children with overweight or obesity: A Randomized Clinical Trial"

### Supplement 3: Additional tables and figures

**eTable 1**. Descriptive characteristics of the ActiveBrains participants meeting the per-protocol criteria at baseline.

|  |  | All | |  | Control group | |  | Exercise group | |
| --- | --- | --- | --- | --- | --- | --- | --- | --- | --- |
|  |  | N | Mean (SD) or % |  | N | Mean (SD) or % |  | N | Mean (SD) or % |
| *Sex* |  |  |  |  |  |  |  |  |  |
| Girls (n %) |  | 41 | 41% |  | 25 | 48% |  | 16 | 39% |
| Boys (n %) |  | 58 | 59% |  | 27 | 52% |  | 31 | 76% |
| At risk of dyslipidemia (n %) |  | 43 | 44% |  | 27 | 52% |  | 16 | 28% |
| Pre-diabetes (n %) |  | 3 | 3% |  | 0 | 0% |  | 3 | 6% |
| Pre-hypertension (n %) |  | 10 | 10% |  | 5 | 10% |  | 5 | 9% |
| Unfit (n %) |  | 62 | 63% |  | 32 | 62% |  | 30 | 53% |
| At risk of metabolic syndrome (n %) |  | 25 | 25% |  | 7 | 13% |  | 18 | 38% |
| At risk of anxiety (n %) |  | 18 | 19% |  | 11 | 22% |  | 7 | 16% |
| At risk of depression (n %) |  | 3 | 3% |  | 1 | 2% |  | 2 | 4% |
| Age (y) |  | 99 | 10.0 (1.1) |  | 51 | 10.1 (1.1) |  | 47 | 10.0 (1.1) |
| Peak height velocity (y) |  | 99 | -2.2 (1.0) |  | 51 | -2.1 (1.1) |  | 47 | -2.4 (0.9) |
| *Cardiometabolic health* |  |  |  |  |  |  |  |  |  |
| LDL (mg/dL) |  | 86 | 100.7 (100.7) |  | 45 | 102.4 (102.4) |  | 41 | 98.9 (98.9) |
| HDL (mg/dL) |  | 95 | 51.0 (51.0) |  | 50 | 49.7 (49.7) |  | 45 | 52.4 (52.4) |
| Triglycerides (mg/dL) |  | 95 | 95.8 (95.8) |  | 50 | 99.0 (99.0) |  | 45 | 92.2 (92.2) |
| Triglycerides-to-HDL (mg/dL) |  | 95 | 2.1 (2.1) |  | 50 | 2.2 (2.2) |  | 45 | 2.0 (2.0) |
| Insulin (mg/dL) |  | 90 | 13.5 (13.5) |  | 47 | 13.2 (13.2) |  | 43 | 13.7 (13.7) |
| Glucose (mg/dL) |  | 96 | 86.3 (86.3) |  | 50 | 84.7 (84.7) |  | 46 | 88.0 (88.0) |
| HOMA |  | 89 | 2.9 (2.9) |  | 46 | 2.8 (2.8) |  | 43 | 3.0 (3.0) |
| Systolic blood pressure (mmHG) |  | 97 | 100.5 (100.5) |  | 50 | 100.1 (100.1) |  | 47 | 101.1 (101.1) |
| Diastolic blood pressure (mmHG) |  | 97 | 57.2 (57.2) |  | 50 | 56.5 (56.5) |  | 47 | 58.0 (58.0) |
| Body mass index (kg/m^2^) |  | 99 | 26.8 (26.8) |  | 52 | 26.3 (26.3) |  | 47 | 27.4 (27.4) |
| Body mass index (WHO z-score) |  | 99 | 3.0 (3.0) |  | 52 | 2.9 (2.9) |  | 47 | 3.1 (3.1) |
| Fat mass index (kg/m^2^) |  | 98 | 11.8 (11.8) |  | 51 | 11.3 (11.3) |  | 47 | 12.4 (12.4) |
| Lean mass index (kg/m^2^) |  | 98 | 13.9 (13.9) |  | 51 | 13.9 (13.9) |  | 47 | 14.0 (14.0) |
| Waist circumference (cm) |  | 99 | 90.5 (90.5) |  | 52 | 89.7 (89.7) |  | 47 | 91.3 (91.3) |
| Visceral adipose tissue (g) |  | 80 | 402.1 (402.1) |  | 42 | 390.4 (390.4) |  | 38 | 415.1 (415.1) |
| CRF performance (laps) |  | 97 | 15.6 (15.6) |  | 50 | 16.1 (16.1) |  | 47 | 15.0 (15.0) |
| CRF (VO_2_max, ml/kg/min) |  | 97 | 40.6 (40.6) |  | 50 | 40.7 (40.7) |  | 47 | 40.4 (40.4) |
| Speed-agility fitness (s) |  | 97 | 15.1 (15.1) |  | 50 | 15.0 (15.0) |  | 47 | 15.3 (15.3) |
| Upper-limb muscular strength (kg) |  | 98 | 16.9 (16.9) |  | 51 | 17.1 (17.1) |  | 47 | 16.7 (16.7) |
| Lower-limb muscular strength (cm) |  | 97 | 104.4 (104.4) |  | 50 | 106.3 (106.3) |  | 47 | 102.3 (102.3) |
| *Children-reported mental health* |  |  |  |  |  |  |  |  |  |
| Stress (0 – 30)* |  | 96 | 5.8 (5.8) |  | 50 | 6.2 (6.2) |  | 46 | 5.4 (5.4) |
| Anxiety (20 – 60)* |  | 95 | 33.6 (33.6) |  | 50 | 34.1 (34.1) |  | 45 | 33.0 (33.0) |
| Depression (0 – 54)* |  | 96 | 8.4 (8.4) |  | 50 | 9.0 (9.0) |  | 46 | 7.8 (7.8) |
| Negative affect (10 – 30)* |  | 95 | 16.0 (16.0) |  | 50 | 16.3 (16.3) |  | 45 | 15.7 (15.7) |
| Positive affect (10 – 30)* |  | 97 | 24.4 (24.4) |  | 51 | 24.5 (24.5) |  | 46 | 24.3 (24.3) |
| Happiness (4 – 28)* |  | 99 | 22.7 (22.7) |  | 52 | 22.4 (22.4) |  | 47 | 23.1 (23.1) |
| Optimism (6 – 30)* |  | 98 | 22.0 (22.0) |  | 51 | 22.1 (22.1) |  | 47 | 22.0 (22.0) |
| Self-efficacy (10 – 40)* |  | 98 | 30.9 (30.9) |  | 51 | 30.5 (30.5) |  | 47 | 31.3 (31.3) |
| Self-concept (30 – 300)* |  | 97 | 227.1 (227.1) |  | 50 | 225.4 (225.4) |  | 47 | 228.8 (228.8) |
| Self-esteem (10 – 40)* |  | 98 | 33.0 (33.0) |  | 51 | 32.6 (32.6) |  | 47 | 33.3 (33.3) |

Data analyses were primarily conducted under the per-protocol principle, i.e., attending to 70% of the sessions or keep the usual lifestyle for exercise and control groups, respectively.

*Score range for the questionnaire.

LDL: low-density lipoprotein; HDL: high-density lipoprotein; HOMA: homeostatic model assessment; CRF: cardiorespiratory fitness; VO_2_max: maximum oxygen consumption, WHO: World Health Organization.

**eTable 2.** Effects of the ActiveBrains exercise program on cardiometabolic health (per-protocol analyses).

|  | Mean (95% CI) | | | | | |  |
| --- | --- | --- | --- | --- | --- | --- | --- |
|  | N_all_ | N | Exercise group* | N | Control group* | Difference between groups | *P* |
| LDL cholesterol (mg/dL) | 71 | 38 |  | 33 |  |  |  |
| Raw score |  |  | 90.35 (85.37 to 95.33) |  | 97.3 (91.95 to 102.64) | -6.95 (-14.27 to 0.37) | 0.063 |
| Z-Score |  |  | -0.42 (-0.62 to -0.22) |  | -0.14 (-0.36 to 0.08) | -0.28 (-0.58 to 0.02) |  |
| HDL cholesterol (mg/dL)^†^ | 79 | 42 |  | 37 |  |  |  |
| Raw score |  |  | 51.57 (48.99 to 54.15) |  | 49.08 (46.33 to 51.83) | 2.49 (-1.3 to 6.27) | 0.195 |
| Z-Score |  |  | 0.04 (-0.2 to 0.27) |  | -0.19 (-0.44 to 0.06) | 0.23 (-0.12 to 0.57) |  |
| Triglycerides (mg/dL) | 86 | 44 |  | 42 |  |  |  |
| Raw score |  |  | 94.15 (83.97 to 104.33) |  | 97.63 (87.21 to 108.05) | -3.48 (-18.06 to 11.09) | 0.636 |
| Z-Score |  |  | -0.05 (-0.27 to 0.17) |  | 0.03 (-0.2 to 0.26) | -0.08 (-0.39 to 0.24) |  |
| TG-to-HDL ratio (mg/dL) | 78 | 41 |  | 37 |  |  |  |
| Raw score |  |  | 1.88 (1.59 to 2.17) |  | 2.12 (1.82 to 2.42) | -0.24 (-0.65 to 0.18) | 0.261 |
| Z-Score |  |  | -0.16 (-0.33 to 0.02) |  | -0.01 (-0.2 to 0.17) | -0.14 (-0.4 to 0.11) |  |
| Insulin (µU/ml) | 76 | 39 |  | 37 |  |  |  |
| Raw score |  |  | 13.36 (11.31 to 15.41) |  | 13.91 (11.8 to 16.02) | -0.55 (-3.49 to 2.39) | 0.710 |
| Z-Score |  |  | -0.01 (-0.25 to 0.22) |  | 0.05 (-0.19 to 0.29) | -0.06 (-0.4 to 0.27) |  |
| Glucose (mg/dL) | 88 | 46 |  | 42 |  |  |  |
| Raw score |  |  | 83.98 (81.59 to 86.38) |  | 83.76 (81.24 to 86.27) | 0.23 (-3.29 to 3.75) | 0.897 |
| Z-Score |  |  | -0.33 (-0.67 to 0.02) |  | -0.36 (-0.72 to 0) | 0.03 (-0.47 to 0.54) |  |
| HOMA | 75 | 39 |  | 36 |  |  |  |
| Raw score |  |  | 2.81 (2.3 to 3.32) |  | 2.93 (2.4 to 3.46) | -0.12 (-0.86 to 0.61) | 0.743 |
| Z-Score |  |  | -0.04 (-0.3 to 0.22) |  | 0.02 (-0.25 to 0.29) | -0.06 (-0.43 to 0.31) |  |
| Systolic and diastolic average (mmHG) | 87 | 46 |  | 41 |  |  |  |
| Raw score |  |  | 78.01 (75.76 to 80.26) |  | 79.88 (77.49 to 82.26) | -1.86 (-5.14 to 1.42) | 0.262 |
| Z-Score |  |  | -0.05 (-0.23 to 0.14) |  | 0.11 (-0.09 to 0.3) | -0.15 (-0.42 to 0.12) |  |
| MAP (mmHG) | 87 | 46 |  | 41 |  |  |  |
| Raw score |  |  | 70.1 (67.93 to 72.27) |  | 71.32 (69.02 to 73.62) | -1.22 (-4.38 to 1.95) | 0.446 |
| Z-Score |  |  | -0.1 (-0.28 to 0.08) |  | 0 (-0.19 to 0.19) | -0.1 (-0.36 to 0.16) |  |
| Body mass index (kg/m^2^) | 92 | 47 |  | 45 |  |  |  |
| Raw score |  |  | 26.34 (26.02 to 26.67) |  | 26.93 (26.59 to 27.26) | -0.59 (-1.06 to -0.12) | 0.011 |
| Z-Score |  |  | -0.13 (-0.23 to -0.04) |  | 0.03 (-0.06 to 0.12) | -0.16 (-0.3 to -0.03) |  |
| WHO z-Score |  |  | -0.69 (-0.75 to -0.63) |  | -0.59 (-0.65 to -0.53) | -0.1 (-0.19 to -0.01) | 0.028 |
| Fat mass index (kg/m^2^) | 92 | 47 |  | 45 |  |  |  |
| Raw score |  |  | 10.75 (10.52 to 10.99) |  | 11.42 (11.18 to 11.66) | -0.67 (-1.01 to -0.33) | 0.0002 |
| Z-Score |  |  | -0.37 (-0.46 to -0.28) |  | -0.1 (-0.2 to -0.01) | -0.27 (-0.4 to -0.14) |  |
| Lean mass index (kg/m^2^) | 92 | 47 |  | 45 |  |  |  |
| Raw score |  |  | 13.58 (13.38 to 13.79) |  | 13.54 (13.33 to 13.75) | 0.05 (-0.25 to 0.34) | 0.757 |
| Z-Score |  |  | -0.24 (-0.38 to -0.1) |  | -0.27 (-0.41 to -0.13) | 0.03 (-0.17 to 0.23) |  |
| Waist circumference (cm) | 92 | 47 |  | 45 |  |  |  |
| Raw score |  |  | 91.85 (90.75 to 92.95) |  | 93.02 (91.89 to 94.14) | -1.17 (-2.74 to 0.41) | 0.145 |
| Z-Score |  |  | 0.14 (0.03 to 0.25) |  | 0.26 (0.14 to 0.37) | -0.12 (-0.28 to 0.04) |  |
| Visceral adipose tissue (g) | 77 | 38 |  | 39 |  |  |  |
| Raw score |  |  | 394.66 (375.13 to 414.19) |  | 426.1 (406.83 to 445.38) | -31.44 (-58.99 to -3.9) | 0.026 |
| Z-Score |  |  | -0.07 (-0.24 to 0.11) |  | 0.21 (0.04 to 0.38) | -0.28 (-0.52 to -0.03) |  |
| CRF performance (laps) | 89 | 46 |  | 43 |  |  |  |
| Raw score |  |  | 18.85 (17.1 to 20.6) |  | 16.09 (14.28 to 17.9) | 2.75 (0.22 to 5.28) | 0.033 |
| Z-Score |  |  | 0.46 (0.22 to 0.71) |  | 0.08 (-0.18 to 0.33) | 0.39 (0.03 to 0.74) |  |
| CRF VO_2_max (ml/kg/min) | 89 | 46 |  | 43 |  |  |  |
| Raw score |  |  | 40.78 (40.16 to 41.4) |  | 39.84 (39.2 to 40.48) | 0.94 (0.05 to 1.84) | 0.039 |
| Z-Score |  |  | 0.08 (-0.16 to 0.31) |  | -0.28 (-0.52 to -0.03) | 0.35 (0.02 to 0.69) |  |
| Speed-agility fitness (sec)‡ | 89 | 46 |  | 43 |  |  |  |
| Raw score |  |  | 14.84 (14.59 to 15.08) |  | 14.96 (14.71 to 15.21) | -0.13 (-0.48 to 0.22) | 0.469 |
| Z-Score |  |  | -0.18 (-0.33 to -0.02) |  | -0.1 (-0.26 to 0.06) | -0.08 (-0.3 to 0.14) |  |
| Upper-limb muscular strength (kg) | 90 | 47 |  | 43 |  |  |  |
| Raw score |  |  | 17.45 (16.93 to 17.97) |  | 17.98 (17.43 to 18.53) | -0.53 (-1.29 to 0.23) | 0.170 |
| Z-Score |  |  | 0.13 (0.01 to 0.26) |  | 0.26 (0.13 to 0.39) | -0.12 (-0.3 to 0.05) |  |
| Lower-limb muscular strength (cm) | 90 | 47 |  | 43 |  |  |  |
| Raw score |  |  | 106.45 (103.14 to 109.77) |  | 109.27 (105.81 to 112.74) | -2.82 (-7.64 to 2) | 0.248 |
| Z-Score |  |  | 0.11 (-0.07 to 0.3) |  | 0.27 (0.08 to 0.46) | -0.16 (-0.42 to 0.11) |  |
| Cardiometabolic risk score 1** | 75 | 41 | -0.163 (-0.405 to 0.079) | 43 | 0.196 (-0.069 to 0.462) | -0.359 (-0.719 to 0.001) | 0.050 |
| Cardiometabolic risk score 2^††^ | 75 | 41 | -0.172 (-0.412 to 0.068) | 43 | 0.208 (-0.056 to 0.472) | -0.38 (-0.738 to -0.021) | 0.038 |

Data analyses were primarily conducted under the per-protocol principle, i.e., attending to 70% of the sessions.

Z-Score values indicate how many standard deviations have the follow-up values changed with respect to the baseline mean and standard deviation. E.g., a 0.50 Z-Score means that the mean value at follow-up is 0.50 standard deviations higher than the mean value at baseline indicating a positive change with negative values indicating the opposite.

*Adjusted for baseline values.

^†^Higher values indicate better health.

^‡^Higher values indicate lower performance.

**Cardiometabolic risk score 1 was calculated as the age- and sex-normalized scores for HDL cholesterol, waist circumference, triglycerides, glucose, and the average of systolic and diastolic blood pressure based on European population reference values.

^††^Cardiometabolic risk score 2 additionally included the CRF.

TG: triglyceride; LDL: low density lipoprotein; HDL: high lipoprotein consumption; MAP: mean arterial pressure; CRF: cardiorespiratory fitness; VO_2_max: maximum oxygen consumption.

**eTable 3.** Effects of the ActiveBrains exercise program on cardiometabolic health (intention-to-treat analyses).

|  | Mean (95% CI) | | | | | | |
| --- | --- | --- | --- | --- | --- | --- | --- |
|  | N_all_ | N | Exercise group* | N | Control group* | Difference between groups | *P* |
| LDL cholesterol (mg/dL) | 109 | 57 |  | 52 |  |  |  |
| Raw score |  |  | 92.914 (88.739 to 97.089) |  | 94.69 (90.319 to 99.062) | -1.776 (-7.829 to 4.276) | 0.562 |
| Z-Score |  |  | -0.32 (-0.487 to -0.153) |  | -0.249 (-0.424 to -0.074) | -0.071 (-0.313 to 0.171) |  |
| HDL cholesterol (mg/dl)† | 109 | 57 |  | 52 |  |  |  |
| Raw score |  |  | 51.981 (49.862 to 54.1) |  | 49.198 (46.979 to 51.417) | 2.783 (-0.291 to 5.857) | 0.034 |
| Z-Score |  |  | 0.132 (-0.066 to 0.33) |  | -0.128 (-0.335 to 0.079) | 0.26 (-0.027 to 0.547) |  |
| Triglycerides (mg/dl) | 109 | 57 |  | 52 |  |  |  |
| Raw score |  |  | 94.085 (85.467 to 102.703) |  | 97.81 (88.786 to 106.835) | -3.725 (-16.225 to 8.775) | 0.556 |
| Z-Score |  |  | -0.043 (-0.234 to 0.149) |  | 0.04 (-0.161 to 0.24) | -0.083 (-0.361 to 0.195) |  |
| TG-to-HDL ratio (mg/dl) | 109 | 57 |  | 52 |  |  |  |
| Raw score |  |  | 1.934 (1.707 to 2.16) |  | 2.124 (1.887 to 2.361) | -0.19 (-0.519 to 0.138) | 0.253 |
| Z-Score |  |  | -0.122 (-0.267 to 0.023) |  | 0 (-0.152 to 0.151) | -0.122 (-0.332 to 0.088) |  |
| Insulin (µU/ml) | 109 | 57 |  | 52 |  |  |  |
| Raw score |  |  | 13.147 (11.681 to 14.614) |  | 13.669 (12.133 to 15.204) | -0.522 (-2.645 to 1.602) | 0.627 |
| Z-Score |  |  | -0.037 (-0.217 to 0.142) |  | 0.026 (-0.161 to 0.214) | -0.064 (-0.323 to 0.196) |  |
| Glucose (mg/dl) | 109 | 57 |  | 52 |  |  |  |
| Raw score |  |  | 84.597 (82.539 to 86.656) |  | 84.041 (81.883 to 86.199) | 0.556 (-2.469 to 3.582) | 0.716 |
| Z-Score |  |  | -0.283 (-0.592 to 0.025) |  | -0.367 (-0.69 to -0.043) | 0.083 (-0.37 to 0.537) |  |
| HOMA | 109 | 57 |  | 52 |  |  |  |
| Raw score |  |  | 2.803 (2.442 to 3.164) |  | 2.894 (2.516 to 3.272) | -0.091 (-0.614 to 0.431) | 0.729 |
| Z-Score |  |  | -0.048 (-0.243 to 0.147) |  | 0.002 (-0.203 to 0.206) | -0.049 (-0.332 to 0.233) |  |
| Systolic and diastolic average (mmHG) | 109 | 57 |  | 52 |  |  |  |
| Raw score |  |  | 78.229 (76.37 to 80.089) |  | 79.859 (77.912 to 81.807) | -1.63 (-4.326 to 1.066) | 0.233 |
| Z-Score |  |  | -0.084 (-0.236 to 0.067) |  | 0.048 (-0.11 to 0.207) | -0.133 (-0.352 to 0.087) |  |
| MAP (mmHG) | 109 | 57 |  | 52 |  |  |  |
| Raw score |  |  | 70.356 (68.535 to 72.177) |  | 71.521 (69.614 to 73.428) | -1.164 (-3.804 to 1.476) | 0.384 |
| Z-Score |  |  | -0.132 (-0.277 to 0.013) |  | -0.039 (-0.191 to 0.112) | -0.093 (-0.302 to 0.117) |  |
| Body mass index (kg/m^2^) | 109 | 57 |  | 52 |  |  |  |
| Raw score |  |  | 26.585 (26.27 to 26.899) |  | 26.903 (26.574 to 27.232) | -0.319 (-0.776 to 0.139) | 0.170 |
| Z-Score |  |  | -0.062 (-0.149 to 0.025) |  | 0.026 (-0.065 to 0.117) | -0.088 (-0.214 to 0.038) |  |
| WHO z-Score |  |  | -0.605 (-0.665 to -0.545) |  | -0.556 (-0.618 to -0.493) | -0.049 (-0.136 to 0.037) | 0.261 |
| Fat mass index (kg/m^2^) | 109 | 57 |  | 52 |  |  |  |
| Raw score |  |  | 13.578 (13.375 to 13.782) |  | 13.66 (13.447 to 13.873) | -0.082 (-0.376 to 0.213) | 0.011 |
| Z-Score |  |  | -0.274 (-0.414 to -0.134) |  | -0.218 (-0.365 to -0.071) | -0.056 (-0.259 to 0.147) |  |
| Lean mass index (kg/m^2^) | 109 | 57 |  | 52 |  |  |  |
| Raw score |  |  | 92.124 (91.128 to 93.119) |  | 92.78 (91.738 to 93.822) | -0.656 (-2.098 to 0.785) | 0.584 |
| Z-Score |  |  | 0.194 (0.094 to 0.295) |  | 0.261 (0.155 to 0.366) | -0.066 (-0.212 to 0.079) |  |
| Waist circumference (cm) | 109 | 57 |  | 52 |  |  |  |
| Raw score |  |  | 397.313 (381.94 to 412.686) |  | 417.309 (401.213 to 433.406) | -19.996 (-42.274 to 2.282) | 0.369 |
| Z-Score |  |  | -0.02 (-0.168 to 0.128) |  | 0.172 (0.017 to 0.327) | -0.193 (-0.407 to 0.022) |  |
| Visceral adipose tissue (g) | 109 | 57 |  | 52 |  |  |  |
| Raw score |  |  | 19.269 (17.434 to 21.103) |  | 17.928 (16.008 to 19.849) | 1.34 (-1.315 to 3.996) | 0.078 |
| Z-Score |  |  | 0.424 (0.185 to 0.662) |  | 0.249 (-0.001 to 0.499) | 0.174 (-0.171 to 0.52) |  |
| CRF performance (laps) | 109 | 57 |  | 52 |  |  |  |
| Raw score |  |  | 40.943 (40.34 to 41.545) |  | 40.277 (39.646 to 40.908) | 0.666 (-0.206 to 1.539) | 0.319 |
| Z-Score |  |  | 0.086 (-0.137 to 0.309) |  | -0.161 (-0.394 to 0.073) | 0.247 (-0.076 to 0.57) |  |
| CRF VO_2_max (ml/kg/min) | 109 | 57 |  | 52 |  |  |  |
| Raw score |  |  | 14.787 (14.552 to 15.023) |  | 14.849 (14.602 to 15.095) | -0.061 (-0.403 to 0.28) | 0.133 |
| Z-Score |  |  | -0.207 (-0.357 to -0.057) |  | -0.168 (-0.325 to -0.01) | -0.039 (-0.257 to 0.179) |  |
| Speed-agility (sec)‡ | 109 | 57 |  | 52 |  |  |  |
| Raw score |  |  | 17.641 (17.155 to 18.127) |  | 18.026 (17.517 to 18.535) | -0.384 (-1.089 to 0.32) | 0.723 |
| Z-Score |  |  | 0.193 (0.076 to 0.31) |  | 0.286 (0.163 to 0.408) | -0.093 (-0.262 to 0.077) |  |
| Upper-limb muscular strength (kg) | 109 | 57 |  | 52 |  |  |  |
| Raw score |  |  | 107.434 (104.459 to 110.41) |  | 108.789 (105.673 to 111.905) | -1.355 (-5.669 to 2.959) | 0.282 |
| Z-Score |  |  | 0.129 (-0.034 to 0.292) |  | 0.204 (0.033 to 0.374) | -0.074 (-0.311 to 0.162) |  |
| Lower-limb muscular strength (cm) | 109 | 57 |  | 52 |  |  |  |
| Raw score |  |  | -0.118 (-0.324 to 0.089) |  | 0.129 (-0.087 to 0.345) | -0.247 (-0.546 to 0.053) | 0.535 |
| Z-Score |  |  | -0.121 (-0.325 to 0.083) |  | 0.133 (-0.081 to 0.346) | -0.254 (-0.55 to 0.042) |  |
| Cardiometabolic risk score 1** | 109 | 57 | 13.578 (13.375 to 13.782) | 52 | 13.66 (13.447 to 13.873) | -0.082 (-0.376 to 0.213) | 0.105 |
| Cardiometabolic risk score 2^††^ | 109 | 57 | -0.274 (-0.414 to -0.134) | 52 | -0.218 (-0.365 to -0.071) | -0.056 (-0.259 to 0.147) | 0.091 |

Data analyses were conducted under the intention-to-treat principle, i.e., including all participants and imputing the missing data using predictive mean matching multiple imputations. Z-score values indicate how many standard deviations have the follow-up values changed with respect to the baseline mean and standard deviation. E.g., a 0.50 Z-score means that the mean value at follow-up is 0.50 standard deviations higher than the mean value at baseline indicating a positive change with negative values indicating the opposite.

*Adjusted for baseline values.

^†^Higher values indicate better health.

^‡^Higher values indicate lower performance

**Cardiometabolic risk score 1 was calculated as the age- and sex-normalized scores for HDL cholesterol, waist circumference, triglycerides, glucose, and the average of systolic and diastolic blood pressure based on European population reference values.

^††^Cardiometabolic risk score 2 additionally included the CRF.

TG: triglyceride; LDL: low density lipoprotein; HDL: high lipoprotein consumption; MAP: mean arterial pressure; CRF: cardiorespiratory fitness; VO_2_max: maximum oxygen consumption.

**eTable 4.** Effects of the ActiveBrains exercise program on mental health (per-protocol analyses).

|  | Mean (95% CI) | | | | | |  |
| --- | --- | --- | --- | --- | --- | --- | --- |
|  | N_all_ | N | Exercise group* | N | Control group* | Difference between groups | *P* |
| *Psychological ill-being†* |  |  |  |  |  |  |  |
| Stress | 88 | 46 |  | 42 |  |  |  |
| Raw score |  |  | 5.17 (4.38 to 5.96) |  | 5.05 (4.22 to 5.88) | 0.12 (-1.04 to 1.27) | 0.842 |
| Z-Score |  |  | -0.2 (-0.44 to 0.04) |  | -0.24 (-0.49 to 0.02) | 0.04 (-0.32 to 0.39) |  |
| Anxiety | 84 | 45 |  | 39 |  |  |  |
| Raw score |  |  | 32.04 (30.16 to 33.92) |  | 30.21 (28.19 to 32.23) | 1.83 (-0.93 to 4.59) | 0.191 |
| Z-Score |  |  | -0.21 (-0.47 to 0.05) |  | -0.46 (-0.74 to -0.18) | 0.25 (-0.13 to 0.63) |  |
| Depression | 88 | 44 |  | 44 |  |  |  |
| Raw score |  |  | 7.31 (5.91 to 8.7) |  | 7.31 (5.91 to 8.7) | 0 (-1.98 to 1.98) | 0.999 |
| Z-Score |  |  | -0.21 (-0.49 to 0.06) |  | -0.21 (-0.49 to 0.06) | 0 (-0.39 to 0.39) |  |
| Negative affect | 80 | 44 |  | 36 |  |  |  |
| Raw score |  |  | 15.71 (14.67 to 16.74) |  | 16.16 (15.02 to 17.31) | -0.45 (-2 to 1.1) | 0.561 |
| Z-Score |  |  | -0.09 (-0.39 to 0.2) |  | 0.04 (-0.29 to 0.36) | -0.13 (-0.57 to 0.31) |  |
| Composite score | 73 | 39 | 0.07 (-0.13 to 0.27) | 34 | -0.07 (-0.28 to 0.15) | 0.14 (-0.15 to 0.43) | 0.343 |
| *Psychological well-being* |  |  |  |  |  |  |  |
| Positive affect | 81 | 44 |  | 37 |  |  |  |
| Raw score |  |  | 24.93 (23.98 to 25.88) |  | 24.94 (23.91 to 25.98) | -0.01 (-1.42 to 1.39) | 0.953 |
| Z-Score |  |  | 0.18 (-0.14 to 0.5) |  | 0.18 (-0.17 to 0.53) | 0 (-0.48 to 0.47) |  |
| Happiness | 92 | 47 |  | 45 |  |  |  |
| Raw score |  |  | 22.79 (21.78 to 23.8) |  | 23.6 (22.56 to 24.63) | -0.81 (-2.26 to 0.64) | 0.304 |
| Z-Score |  |  | 0.01 (-0.24 to 0.27) |  | 0.22 (-0.04 to 0.48) | -0.21 (-0.58 to 0.16) |  |
| Optimism | 87 | 46 |  | 41 |  |  |  |
| Raw score |  |  | 22.56 (21.44 to 23.68) |  | 22.32 (21.14 to 23.51) | 0.24 (-1.39 to 1.86) | 0.645 |
| Z-Score |  |  | 0.13 (-0.15 to 0.41) |  | 0.07 (-0.22 to 0.37) | 0.06 (-0.34 to 0.46) |  |
| Self-efficacy | 90 | 47 |  | 43 |  |  |  |
| Raw score |  |  | 31.05 (29.7 to 32.41) |  | 31.61 (30.2 to 33.03) | -0.56 (-2.52 to 1.4) | 0.651 |
| Z-Score |  |  | 0.04 (-0.23 to 0.32) |  | 0.15 (-0.13 to 0.44) | -0.11 (-0.51 to 0.29) |  |
| Self-concept | 79 | 43 |  | 36 |  |  |  |
| Raw score |  |  | 219.53 (211.27 to 227.78) |  | 223.15 (214.12 to 232.17) | -3.62 (-15.86 to 8.62) | 0.557 |
| Z-Score |  |  | -0.26 (-0.55 to 0.02) |  | -0.14 (-0.45 to 0.18) | -0.13 (-0.55 to 0.3) |  |
| Self-esteem | 87 | 47 |  | 40 |  |  |  |
| Raw score |  |  | 35.17 (33.99 to 36.35) |  | 34.8 (33.52 to 36.08) | 0.37 (-1.37 to 2.11) | 0.609 |
| Z-Score |  |  | 0.47 (0.22 to 0.72) |  | 0.39 (0.12 to 0.66) | 0.08 (-0.29 to 0.45) |  |
| Composite score | 72 | 41 | -0.05 (-0.3 to 0.2) | 31 | 0.08 (-0.21 to 0.36) | -0.13 (-0.51 to 0.25) | 0.503 |
| *Total mental health* |  |  |  |  |  |  |  |
| Composite score | 64 | 36 | -0.12 (-0.36 to 0.12) | 28 | 0.11 (-0.16 to 0.39) | -0.24 (-0.6 to 0.13) | 0.206 |

Data analyses were primarily conducted under the per-protocol principle, i.e., attending to 70% of the sessions. Z-score values indicate how many standard deviations have the follow-up values changed with respect to the baseline mean and standard deviation. E.g., a 0.50 Z-score means that the mean value at follow-up is 0.50 standard deviations higher than the mean value at baseline, indicating a positive change with negative values indicating the opposite.

*Adjusted for baseline values.

^†^Higher values indicate lower mental health.

**eTable 5.** Effects of the ActiveBrains exercise program on mental health (intention-to-treat analyses).

|  | Mean (95% CI) | | | | | |  |
| --- | --- | --- | --- | --- | --- | --- | --- |
|  | N_all_ | N | Exercise group* | N | Control group* | Difference between groups | P |
| *Psychological ill-being*† |  |  |  |  |  |  |  |
| Stress | 109 | 57 |  | 52 |  |  |  |
| Raw score |  |  | 5.24 (4.49 to 5.98) |  | 5.13 (4.34 to 5.92) | 0.11 (-0.98 to 1.20) | 0.845 |
| Z-Score |  |  | -0.16 (-0.4 to 0.08) |  | -0.19 (-0.45 to 0.06) | 0.03 (-0.32 to 0.39) |  |
| Anxiety | 109 | 57 |  | 52 |  |  |  |
| Raw score |  |  | 32.11 (30.53 to 33.68) |  | 30.94 (29.27 to 32.61) | 1.17 (-1.13 to 3.46) | 0.316 |
| Z-Score |  |  | -0.2 (-0.41 to 0.02) |  | -0.36 (-0.59 to -0.13) | 0.16 (-0.16 to 0.48) |  |
| Depression | 109 | 57 |  | 52 |  |  |  |
| Raw score |  |  | 7.42 (6.19 to 8.66) |  | 7.8 (6.49 to 9.1) | -0.37 (-2.18 to 1.43) | 0.683 |
| Z-Score |  |  | -0.19 (-0.42 to 0.05) |  | -0.12 (-0.37 to 0.13) | -0.07 (-0.41 to 0.27) |  |
| Negative affect | 109 | 57 |  | 52 |  |  |  |
| Raw score |  |  | 15.72 (14.87 to 16.58) |  | 16.33 (15.42 to 17.23) | -0.61 (-1.85 to 0.64) | 0.337 |
| Z-Score |  |  | -0.14 (-0.39 to 0.12) |  | 0.04 (-0.23 to 0.31) | -0.18 (-0.55 to 0.19) |  |
| Composite score | 109 | 57 | -0.02 (-0.22 to 0.18) | 52 | 0.02 (-0.19 to 0.23) | -0.03 (-0.33 to 0.26) | 0.814 |
| *Psychological well-being* |  |  |  |  |  |  |  |
| Positive affect | 109 | 57 |  | 52 |  |  |  |
| Raw score |  |  | 24.71 (23.83 to 25.59) |  | 24.7 (23.77 to 25.63) | 0.01 (-1.27 to 1.29) | 0.984 |
| Z-Score |  |  | 0.09 (-0.21 to 0.38) |  | 0.08 (-0.23 to 0.39) | 0 (-0.42 to 0.43) |  |
| Happiness | 109 | 57 |  | 52 |  |  |  |
| Raw score |  |  | 23.01 (22.1 to 23.92) |  | 23.1 (22.14 to 24.06) | -0.09 (-1.42 to 1.24) | 0.892 |
| Z-Score |  |  | 0.07 (-0.16 to 0.29) |  | 0.09 (-0.15 to 0.33) | -0.02 (-0.36 to 0.31) |  |
| Optimism | 109 | 57 |  | 52 |  |  |  |
| Raw score |  |  | 22.73 (21.73 to 23.74) |  | 21.9 (20.83 to 22.97) | 0.83 (-0.63 to 2.30) | 0.263 |
| Z-Score |  |  | 0.21 (-0.05 to 0.46) |  | -0.01 (-0.28 to 0.27) | 0.21 (-0.16 to 0.58) |  |
| Self-efficacy | 109 | 57 |  | 52 |  |  |  |
| Raw score |  |  | 31.47 (30.29 to 32.64) |  | 31.62 (30.38 to 32.87) | -0.16 (-1.88 to 1.56) | 0.857 |
| Z-Score |  |  | 0.11 (-0.13 to 0.36) |  | 0.15 (-0.11 to 0.41) | -0.03 (-0.39 to 0.33) |  |
| Self-concept | 109 | 57 |  | 52 |  |  |  |
| Raw score |  |  | 224.4 (218.94 to 229.86) |  | 221.92 (216.14 to 227.69) | 2.48 (-5.48 to 10.44) | 0.538 |
| Z-Score |  |  | -0.07 (-0.26 to 0.12) |  | -0.16 (-0.36 to 0.04) | 0.09 (-0.19 to 0.36) |  |
| Self-esteem | 109 | 57 |  | 52 |  |  |  |
| Raw score |  |  | 35.14 (34.03 to 36.25) |  | 34.15 (32.98 to 35.32) | 0.99 (-0.63 to 2.61) | 0.227 |
| Z-Score |  |  | 0.46 (0.22 to 0.69) |  | 0.25 (-0.002 to 0.50) | 0.21 (-0.13 to 0.55) |  |
| Composite score | 109 | 57 | 0.04 (-0.19 to 0.26) | 52 | -0.04 (-0.27 to 0.2) | 0.07 (-0.25 to 0.4) | 0.654 |
| *Total mental health* |  |  |  |  |  |  |  |
| Composite score | 109 | 57 | 0.02 (-0.19 to 0.23) | 52 | -0.02 (-0.24 to 0.2) | 0.04 (-0.27 to 0.34) | 0.816 |

Data analyses were conducted under the intention-to-treat principle, i.e., including all participants and imputing the missing data using predictive mean matching multiple imputations. Z-score values indicate how many standard deviations have the follow-up values changed with respect to the baseline mean and standard deviation. E.g., a 0.50 Z-score means that the mean value at follow-up is 0.50 standard deviations higher than the mean value at baseline indicating a positive change, with negative values indicating the opposite.

*Adjusted for baseline values.

^†^Higher values indicate lower mental health.

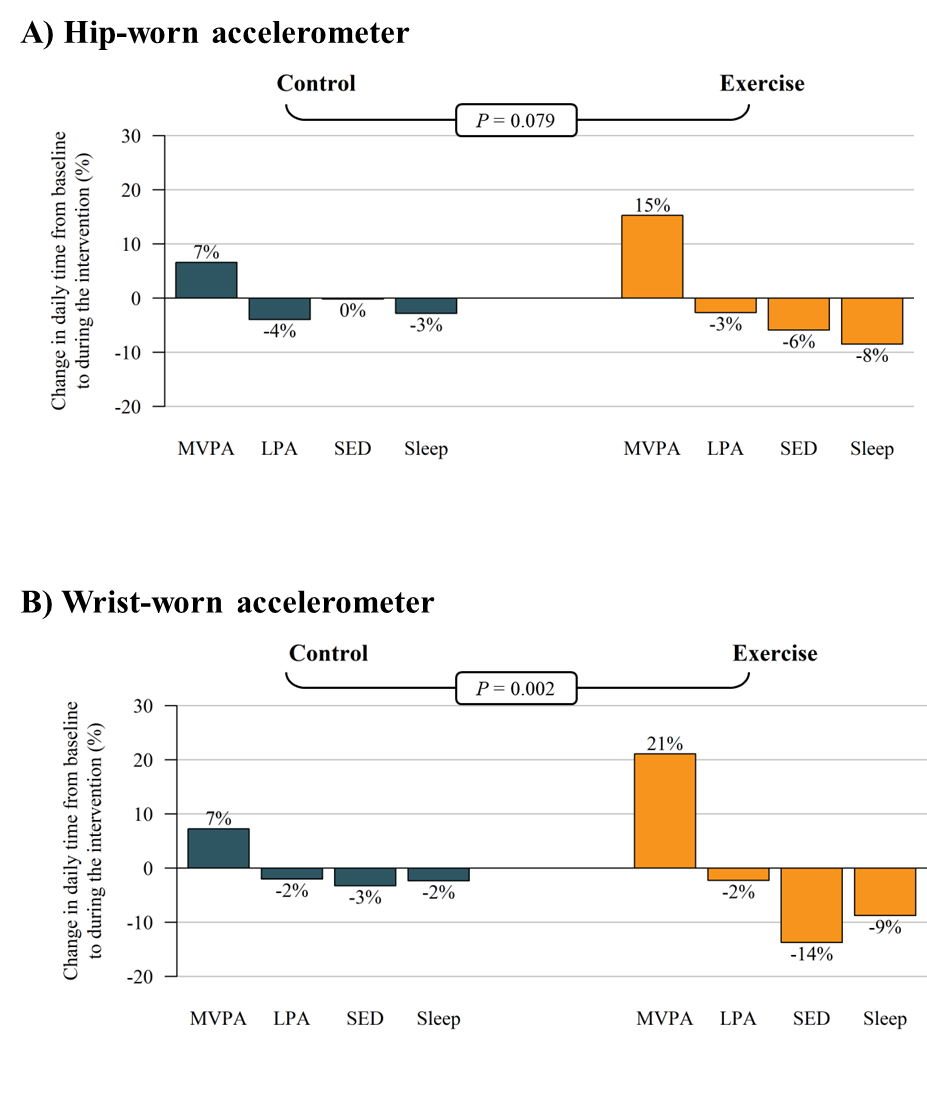

**eFigure 1.** Change in daily activity composition induced by the exercise program as measured with hip- (panel A) and wrist-worn (panel B) accelerometers.

Data analyses were primarily conducted under the per-protocol principle, i.e., attending to 70% of the sessions. Isometric log-ratios between each group’s compositional mean and the overall compositional mean after centering the data at baseline and during exercise were calculated.

*P* value from Hotelling’s T-squared test for pair-wise comparison of multivariate means.

MVPA: moderate-to-vigorous physical activity; LPA: light physical activity; SED: sedentary time.
